# An evaluation of the early impact of the COVID-19 pandemic on Zambia’s routine immunization program

**DOI:** 10.1101/2022.08.12.22278710

**Authors:** Amy K. Winter, Saki Takahashi, Andrea Carcelén, Kyla Hayford, Wilbroad Mutale, Francis Mwansa, Nyambe Sinyange, David Ngula, William J. Moss, Simon Mutembo

**Affiliations:** Department of Epidemiology and Biostatistics, University of Georgia, Athens, GA, USA; Division of HIV, Infectious Diseases & Global Medicine, University of California, San Francisco, CA, USA; Department of International Health, International Vaccine Access Center, Johns Hopkins Bloomberg School of Public Health, Baltimore, MD, USA; School of Public Health, University of Zambia, Lusaka, Zambia; Child Health Unit, Directorate of Public Health and Research, Ministry of Health, Lusaka, Zambia; Field Epidemiology Training Program, Zambia National Public Health Institute, Lusaka, Zambia; Department of Epidemiology, Johns Hopkins Bloomberg School of Public Health, Baltimore, MD, USA

## Abstract

**Background:** Implications of the COVID-19 pandemic for both populations and healthcare systems are vast. In addition to morbidity and mortality from COVID-19, the pandemic also has disrupted local health systems, including reductions or delays in routine vaccination services and catch-up vaccination campaigns that could lead to outbreaks of other infectious diseases that result in an additional burden of disease and strain on the healthcare system.

**Methods and Findings:** We evaluated the impact of the COVID-19 pandemic on Zambia’s routine childhood immunization program in 2020 using multiple sources of data. We relied on district-level administrative vaccination coverage data and Zambia’s 2018 Demographic and Health Survey to project disruptions to routine childhood vaccination within the pandemic year 2020 (N=5,670). Next, we leveraged serological data to predict age-specific measles seroprevalence and assessed the impact of changes in vaccination coverage on measles outbreak risk in each district. We found minor disruptions to routine administration of measles-rubella and pentavalent vaccines in 2020. This was in part due to Zambia’s Child Health Week held in June of 2020 which helped to reach children missed during the first six months of the year. We estimated that the two-month delay in a measles-rubella vaccination campaign, originally planned for September of 2020 but conducted in November of 2020 as a result of the pandemic, had little impact on modeled district-specific measles outbreak risks.

**Conclusions:** The pandemic only minimally increased the number of children missed by measles-rubella and pentavalent vaccines in 2020. However, the ongoing SARS-CoV-2 transmission since our analysis concluded means efforts to maintain routine immunization services and minimize the risk of measles outbreaks will continue to be critical. Fortunately, the methodological framework developed in this analysis relied on routinely collected data and can be used to evaluate COVID-19 pandemic disruptions in Zambia following 2020 and in other countries or for other vaccines at a sub-national level.

## Introduction

SARS-CoV-2 has been responsible for hundreds of millions of confirmed cases of COVID-19 globally (1). Implications of the pandemic for both the population and the healthcare system are vast. In addition to direct morbidity and mortality from COVID-19, the pandemic also has the potential to cause disruptions to local health systems. Disruptions of particular concern are to routine immunization programs used to control the spread of other infectious diseases (2).

Disruptions to immunization programs could lead to outbreaks of vaccine-preventable diseases and further complicate responses to the COVID-19 pandemic (3). Reasons for disruptions include supply-side issues, including international and domestic supply chain disruptions, border closures and trade restrictions, and assignment of vaccination staff to COVID-19 control activities (4). Additionally, demand-side related issues may also negatively impact uptake of vaccines such as maternal reluctance to seek vaccinations for their children because they do not want to risk being exposed to SARS-CoV-2 at health facilities when they take their children for vaccinations.

As governments across the world try to control the pandemic by implementing population wide lock downs, halting mass gatherings, and closing borders, routine immunization program have been sliding backwards (2). According to the World Health Organization (WHO) the suspension of vaccination services in over 68 countries during the early stages of the pandemic put over 80 million infants younger than one year of age at risk of measles (5). With approximately 90 mass vaccination campaigns postponed in 2020, 24 million children who were to benefit from these campaigns are at risk of being unvaccinated against measles and rubella (6). As a result, vaccination coverage has declined and increased cases of polio and diphtheria have been reported, for example, in Pakistan (7).

We evaluated the impact of Zambia’s COVID-19 outbreak on its vaccination program at the sub-national level (i.e., 2nd administrative level). As it is difficult to establish a causal link between the pandemic and vaccination programs, here we compared pre-pandemic and pandemic vaccination program performance and attributed the difference to the pandemic. Findings from this analysis were used by the Zambian Ministry of Health to inform country-specific 2020 vaccination strategic responses in light of the COVID-19 pandemic.

## Methods

We evaluated changes in administration of routine bivalent measles-rubella vaccine dose 1 (MR1) and the pentavalent diphtheria, pertussis, tetanus toxoid, hepatitis B and *Haemophilus influenzae* type b vaccine doses 1 and 3 (Penta1, Penta3). In Zambia, the recommended childhood vaccination schedule includes three Penta doses at 6, 10, and 14 weeks of life and two MR doses at 9 and 18 months of age. We evaluated disruptions to measles-containing vaccines because clusters of few susceptible individuals pose a high risk of measles outbreak. We additionally evaluated coverage of the pentavalent vaccine given that coverage of pentavalent vaccines are used as an indicator of health care system performance (8).

To understand how COVID-19 related health care disruptions potentially increased the risk of vaccine-preventable diseases, we estimated the spatial distribution of unvaccinated children before and during each month of disruptions in a time frame that encompassed two waves of COVID-19 waves in July 2020 and January 2021. We established a baseline vaccination rate and coverage across ages at the district level, estimated the impact of the pandemic as a percent reduction in the rate of vaccination, and finally calculated the number of children missed for each month of disruption. For measles, given the particularly high transmissibility among susceptible individuals, we took one step further and explored measles outbreak risk with and without vaccine disruptions, and evaluated the impact of a two month delay of the national MR vaccination campaign. See Figure 1 for a methodological roadmap summarizing all steps.

**Figure 1:**
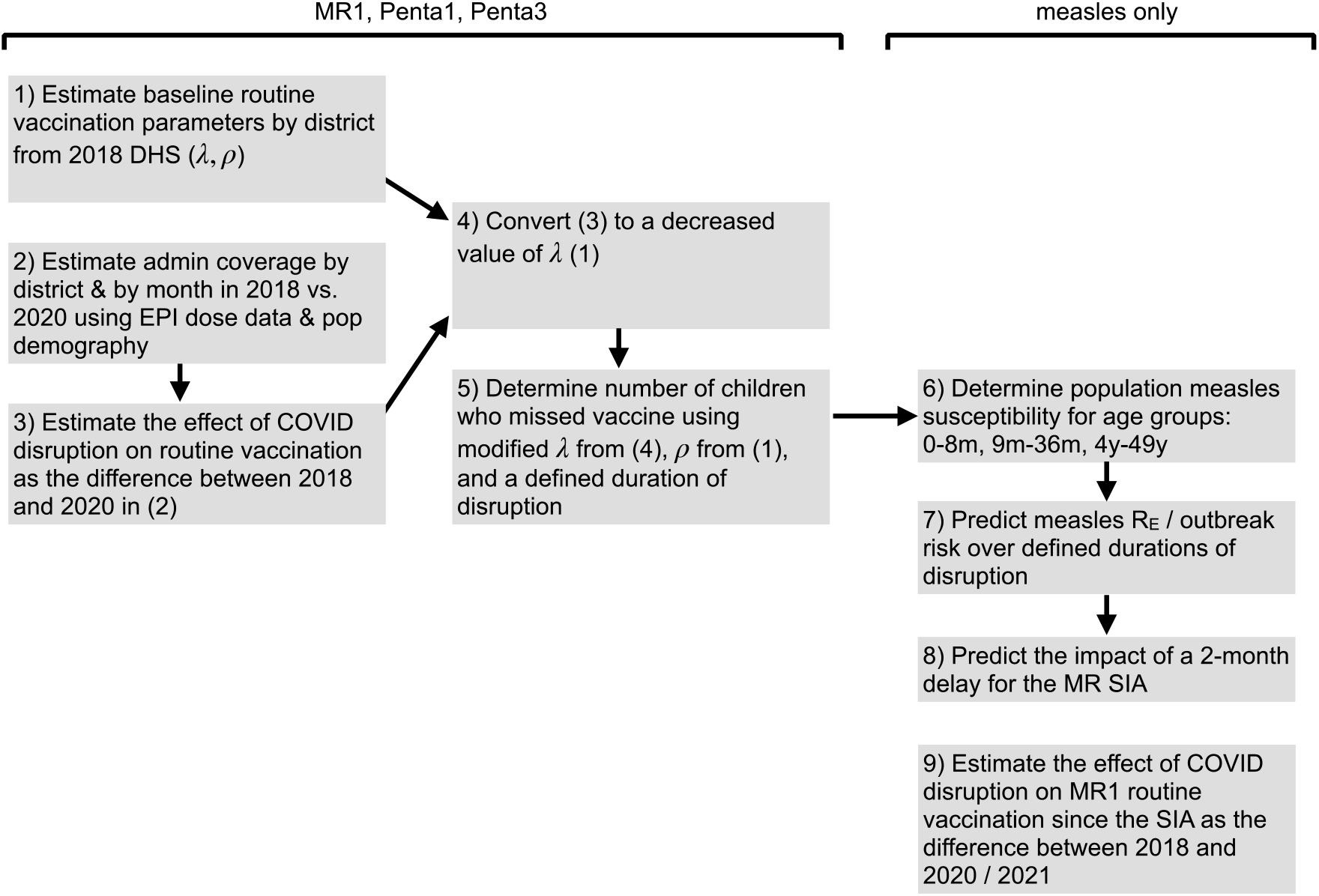
Methodological roadmap.

### Pre-COVID-19 pandemic routine “baseline” vaccination coverage and rates

We relied on childhood vaccination coverage data from Zambia’s 2018 cross-sectional Demographic and Health Survey (DHS) made publicly available by ICF International to estimate routine vaccination coverage by age and by district in Zambia. For each child, we extracted from the DHS: age at the time of survey, whether the child had ever received an MR1, Penta1, and Penta3 vaccine dose (based on vaccination card or report of the parent or guardian), and age at the time of vaccination (if a vaccination card was available). Data were available for 111 of the 115 designated districts in 2018 and for 5,670 children between the ages of 0 and 36 months. We conducted a modified survival analysis accounting for uncensored (i.e. child with vaccination date on vaccination card), left censored (i.e., mother reported vaccination but date of vaccination unknown), and right censored (i.e., child unvaccinated at the time of the DHS survey) data on the age of vaccination, extending a statistical approach previously described (9). The probability that an individual was vaccinated with each vaccine by a given age depends on two estimated parameters: district-specific lifetime probability of being vaccinated through routine vaccination services (*ρ*_*i*_) and the district-specific rate of receiving the vaccine through routine vaccination (*λ*_*i*_) (Fig. S1); the latter accounts for differences in the timeliness of receiving routine vaccination as a function of age. To estimate these district-specific parameters, we assumed they were multivariate normally distributed with a district-specific mean that was specified by a conditional autoregressive model. We extended the granularity of DHS provincially representative data using spatial models that account for the data structure. As a result, district estimates depended on assumptions inherent in the conditional autoregressive model. See Appendix S1for more details.

### Magnitude of disruption from the COVID-19 pandemic

Administrative data of the number of vaccine doses delivered per month in each district from January 2018 to August 2020 for MR1, Penta1, and Penta3 obtained from the Zambia Expanded Programme on Immunization (EPI) were used to estimate disruptions to routine vaccinations due to the COVID-19 pandemic. We estimated a national reduction in the rate of routine vaccination because the administrative vaccination data were not sufficient to evaluate sub-national disruptions to routine vaccination. Given the seasonal nature of vaccine delivery due to Child Health Weeks (Fig. S2), we compared the number of vaccine doses delivered during each month in 2018 and 2020. We selected 2018 as the baseline year for consistency with analysis on pre-COVID-19 vaccination coverage (i.e., DHS data).

We fit a binomial model to each year of data (2018 and 2020), *N*_*it*_ *∼ Binomial (B*_*i*_, *γ*_*t*_), where *N*_*it*_ *i*s the number of doses administered in district *i* and month *t, B*_*i*_ *i*s the size of the birth cohort in district *i*, and *γ*_*t*_ *i*s the estimated proportion of individuals vaccinated in month *t*. We estimated the size of the birth cohort in 2018 and 2020 for each district by *B*_*i*_ *=P*_*i*_ *bk*, where P_*i*_ *i*s the size of the population in the district *i* estimated by aggregating WorldPop population estimates in 10×10 km grid cells over district polygons (10,11), and *bk i*s the proportion of the population who are newborns (age 0) for the respective province *k* in which district *i* is located based on Zambia’s central statistical office projections.

The disruption rate per month was estimated as the percent reduction between the 2018 and 2020 in monthly estimates of *γt*. We incorporated uncertainty by evaluating the percent reduction between upper and lower 95% credible intervals in 2018 and 2020. For example, the lower range of the disruption rate per month was estimated as the percent difference between the 2.5th quantile of the 2018 month estimates and the 97.5th quantile of the 2020 estimates; the upper range of disruption rate per month was estimated as the mean percent difference between the 97.5th quantile of the 2018 month estimates and the 2.5th quantile of the 2020 estimates. Given monthly variation, we calculated the national mean across 10 months to obtain an overall percent reduction in routine rate and range for each vaccination dose, and assumed it was constant into the future.

### Number of children missed by vaccination

We directly applied the national mean percent reduction in the rate of routine vaccination to the estimated district-specific baseline rate of routine vaccinations (*λ*_*i*_) to estimate a modified district-specific proportion vaccinated over age in months. The duration of disruption determined the duration of time individuals were exposed to the reduced rate of vaccination compared to the baseline rate of vaccination. Therefore, for each month of disruption, we estimated a district and age-specific proportion vaccinated between 9 and 36 months of age. The number of children not vaccinated in each district was calculated as the sum of the number of individuals 9 to 36 months old times the proportion unvaccinated between 9 and 36 months (i.e., 1 - proportion vaccinated). Individuals who aged out of this age group and remained unvaccinated were not included in these totals. These estimates were aggregated across districts to obtain national estimates.

### Measles outbreak risk

We evaluated the district-specific additional risk of a measles outbreak for each month of disruption in 2020 and the impact of a national delay in the vaccination campaign from September to November 2020. We focused on measles given its high transmissibility and herd immunity threshold. To estimate outbreak risk, we first needed to estimate age-specific susceptibility. Population susceptibility was determined separately for three age bins: birth to 9 months, 9 months to 36 months (i.e., the age range of individuals with relevant measles vaccination data in the DHS survey), and 36 months to 49 years. We assumed that all infants from birth to nine months old had a level of protection by maternally derived antibodies beginning with 100% at birth and dropping by a rate of 0.45 until 8 months of age, and are susceptible thereafter until vaccinated (12). We assumed no immunity from natural infection given the small number of measles cases reported since 2016 (average of 11 annual cases between 2016-2019) (13).

We estimated susceptibility of individuals between 9 months and 36 months old by applying an age-specific vaccine effectiveness rate to the disruption modified district-specific probability an individual was vaccinated by age in months (as estimated above). We assumed all changes in the proportion susceptible by month of disruption was constrained to the 9 to 36 month old age group who were eligible for MR1 and receiving it at a reduced rate, thereby ignoring the potential role that natural infection may have to reduce the impact of susceptibility on individuals younger than 9 months or older than 36 months. This assumption is justified by the lack of major measles outbreaks in 2020 in Zambia. The febrile-rash surveillance system in Zambia reported only 69 measles cases in 2020 to the World Health Organization, similar to the previous six years (14).

To estimate susceptibility among individuals older than 36 months, we relied on a hierarchical spatial model fit to measles seroprevalence data collected in 2016 (15). Serological data provides the most direct estimate of measles immunity, obtained through vaccination or natural infection (16). The measles IgG 2016 serological data came from a nested serosurvey within the Zambia Population-Based HIV Impact Assessment consisting of 9,852 blood samples collected from individuals one month to 49 years old across all 72 districts (15). The model relied on routinely collected epidemiological data (i.e., vaccination coverage, suspected case data) and demographic data (i.e., age, district, province) to explain the variation in the cross-sectional 2016 seroprevalence data. To project measles seroprevalence in subsequent years, selected covariates from those years were combined with posterior estimates of model parameters. We did not have data on individuals over 49 years of age. We do not have estimates on susceptibility among populations over 49 years of age. However, given these birth cohorts were children when measles virus transmission was endemic in Zambia, it is reasonable to assume they would have been naturally exposed to measles virus and would therefore not be susceptible (17). See Appendix S3 and Figures S3-S6 for more details.

Lastly, to estimate outbreak risk, we calculated the measles effective reproductive number (Re, i.e., the average number of individuals an infectious individual will infect in a partially susceptible population) for each month and district taking into account age-specific susceptibility and age-assortative mixing patterns (18). See Appendix S2 for more details.

### Ethics Statement

For the 2016 measles seroprevalence data, participants provided written informed consent, parental permission was obtained children under 18 years old, and assent was obtained for participants 10-17 years old. This serosurvey was conducted in accordance with relevant guidelines and regulations. Ethical approvals for protocols were provided by Johns Hopkins Bloomberg School of Public Health (00008423) as well as the Tropical Disease Research Center and the National Health Regulatory Agency in Zambia (TDRC/C4/01/2019).

## Results

In our baseline estimates of routine vaccination coverage, we identified variation in district-level estimates of the lifetime probability and monthly rate of being vaccinated (Fig. 2). The median lifetime probability of receiving MR1, Penta1, and Penta3 across districts was 0.937 (range 0.670 to 0.983), 0.990 (range 0.809 to 0.997), and 0.949 (range 0.565 to 0.993), respectively (Fig. 2A-C). There was some consistency, however, in the estimated lifetime probability of being vaccinated across the three vaccines, i.e., districts with relatively lower lifetime probability of vaccination with MR1 also had lower probabilities of vaccination with Penta1 and Penta3 vaccines compared to other districts (correlation MR1 and Penta1 = 0.834; correlation MR1 and Penta3 = 0.764; correlation Penta1 and Penta3 = 0.801). The median monthly rate of routine vaccination for MR1, Penta1, and Penta3 across districts was 0.510 (range 0.287 to 0.730), 0.645 (range 0.376 to 0.862), and 0.412 (range 0.255 to 0.601), respectively (Fig. 2D-F); this is equivalent to a median average age of vaccination (among those vaccinated) of 9.96 months (range 9.34 to 11.49), 1.95 months (range 1.56 to 3.06), and 4.63 months (range 3.86 to 6.12) (Fig. S7). Districts with a lower rate of vaccination for MR1 did not necessarily have a lower rate of vaccination for Penta1 and Penta3 (correlation between MR1 and Penta1 = 0.423, correlation between MR1 and Penta3 = 0.428), although the rates at which Penta1 and Penta3 were administered was correlated (correlation between Penta1 and Penta3 = 0.731). The resulting median and 95% credible intervals in the estimated proportion vaccinated over age in months are displayed in Figures S8-S10 and show good model fit to the raw DHS data.

**Figure 2:**
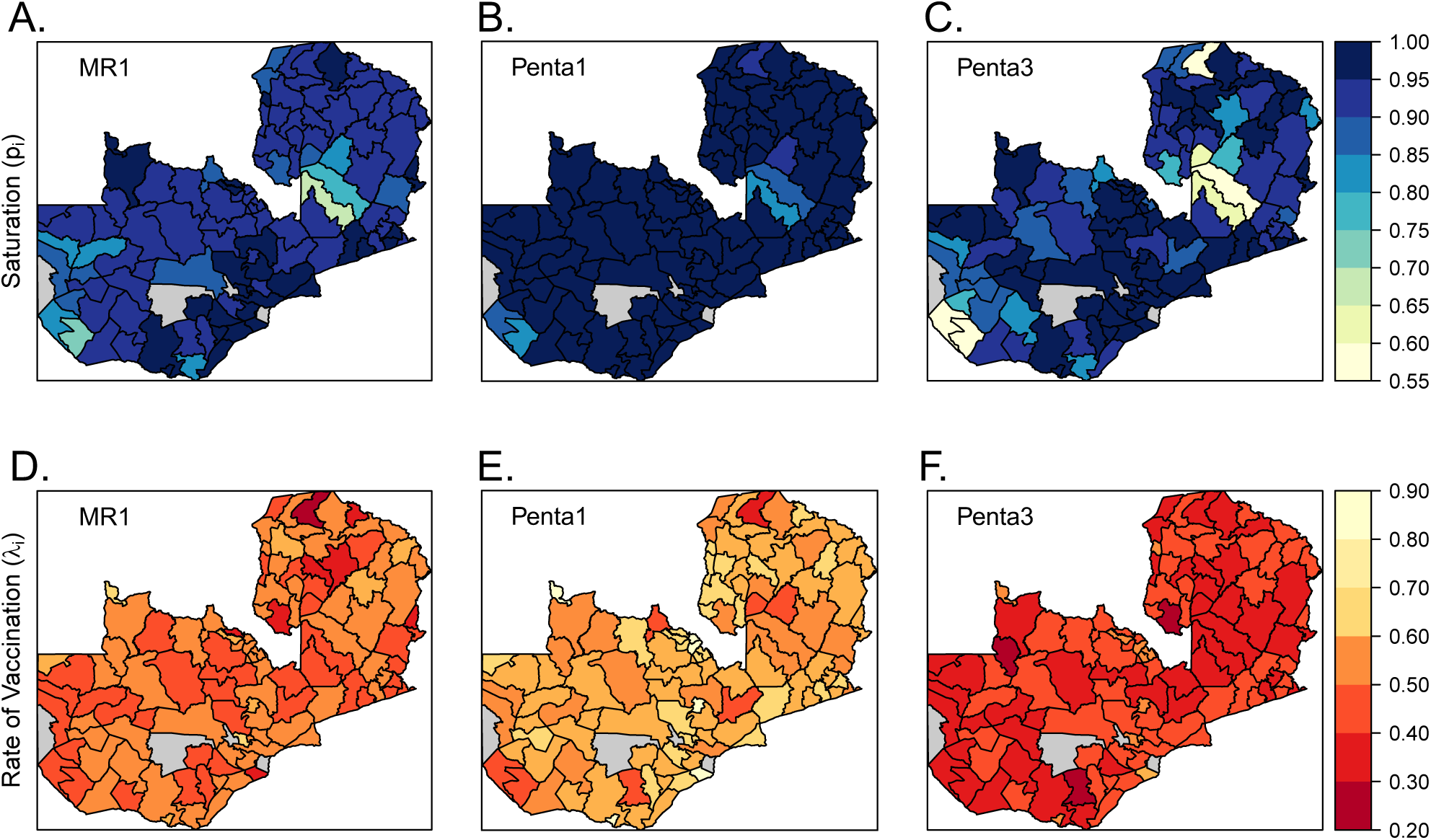
District-level (111 / 115 districts) median parameter estimates of the lifetime probability of being vaccinated via routine vaccination through 36 months of age (saturation parameter) (A-C), and monthly rate of receiving routine vaccination (D-F) for MR1 (A & F), Penta1 (B & E), and Penta 3 (C & F) vaccine doses. There are no parameter estimates for four districts colored in grey due to the lack of any DHS sampling clusters in these districts.

For all three vaccines, we found an increase in the proportion of the annual birth cohort vaccinated in the month of June because of Zambia’s Child Health Week (Fig. 3). The estimated rate of disruptions in routine vaccination due to the COVID-19 pandemic varied between MR1, Penta1, and Penta3. Routine vaccination with MR1 in 2020 was lower from January through May compared to in 2018; however, the Child Health Week in June helped to catch-up some missed children. Routine vaccination with Penta1 and Penta3 had fewer disruptions in January and February 2020 but disruptions were observed from March to June. Similar to MR1, we noted a slight catch-up of missed children in June 2020 for Penta3 vaccination. Routine vaccination with MR1 had the largest difference between 2018 and 2020, with a percent reduction in the mean 2020 estimates from the mean 2018 estimates ranging from a 14.4% reduction to -4.0% reduction (i.e., an increase by 4% in June 2020 compared to June 2018). The mean percent reduction across months was 5.7% (lower bound: 4.1%, upper bound: 7.3%). The percent reduction in routine vaccination in the mean 2020 estimates from the mean 2018 estimates for Penta1 ranged from 9.7% to -1.5% and the mean percent reduction across months was 3.6% (lower bound: 2.0%, upper bound: 5.1%). The percent reduction in routine vaccination in the mean 2020 estimates from the mean 2018 estimates for Penta3 ranged from 11.0% to -3.9% and the mean percent reduction across months was 3.1% (lower bound: 1.4%, upper bound: 4.7%).

**Figure 3:**
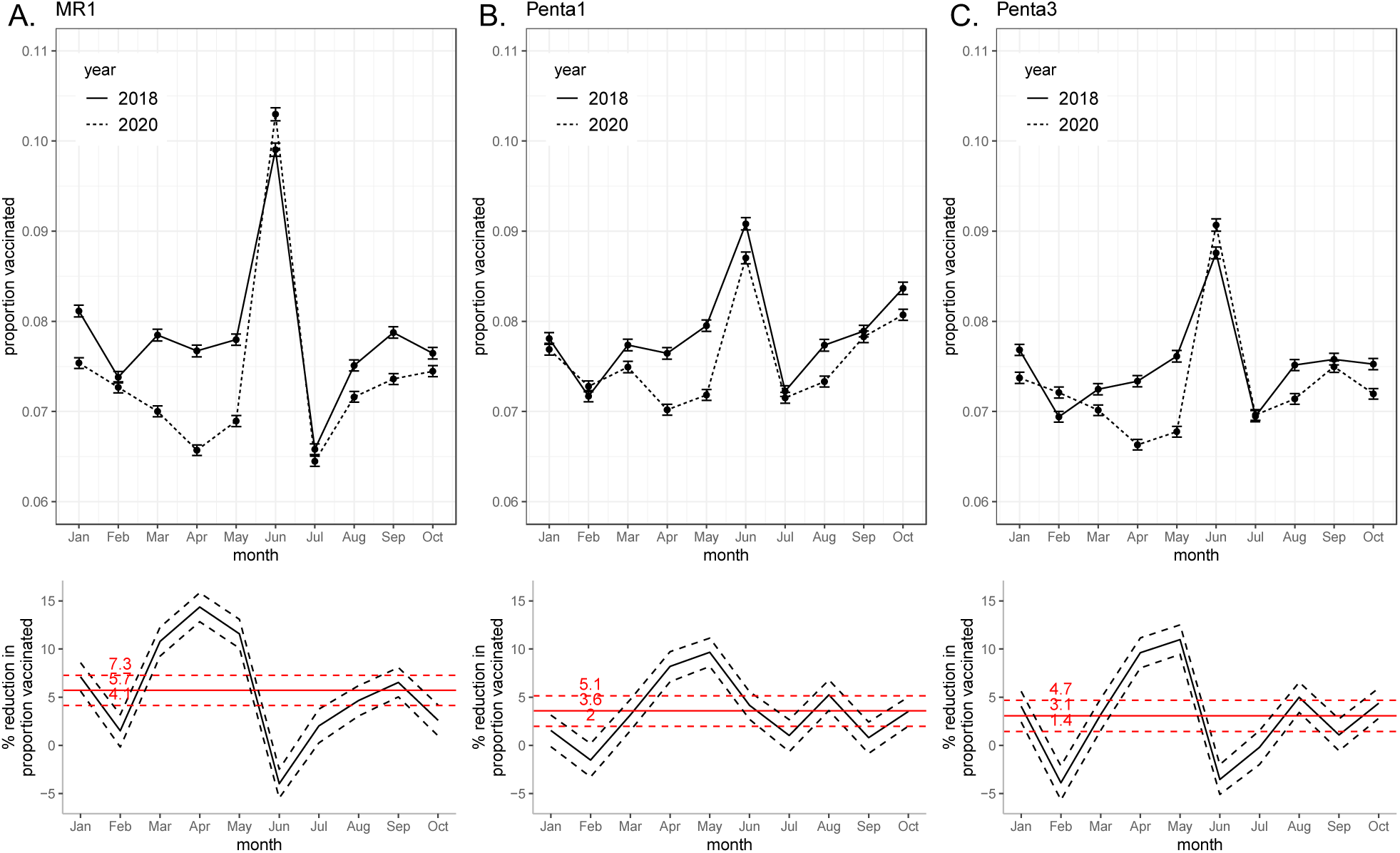
Disruption to routine vaccination for MR1 (A), Penta1 (B), Penta3 (C) based on EPI data. Top row is the proportion of the birth cohort vaccinated January to June in years 2018 and 2020 (mean and 95% credible intervals represented by points and error bars). Bottom row is the percent reduction in proportion vaccinated each month (black lines) and mean across months (red lines); mean (1 - (2020 mean / 2018 mean)), lower bound (1 - (2020 upper bound of 95% credible interval / 2018 lower bound of 95% credible interval)), and upper bound (1 - (2020 lower bound of 95% credible interval / 2018 upper bound of 95% credible interval)).

The number of additional children missed by vaccination in 2020 was minimal compared to the total number missed in our assumed baseline year of 2018. In the reference year of 2018 (i.e., without disruptions from the COVID-19 pandemic, we estimate that 224,522 (95% CI 148,260 - 352,653) children missed MR1, 142,787 (95% CI 106,034 - 228,038) missed Penta1, and 267,250 (95% CI 178,946 - 416,484) missed Penta3 (Fig. 4A-C, Fig. S11). Given the median number of missed vaccinations in a non-disruption year, the percent increase in the number of doses missed in 2020 was largest for MR1 (2.5%) followed by Penta1 (2.1%) and Penta3 (1.4%). An additional 5,680 (95% CI 4,019 - 7,395) children missed MR1 after 10 months of disruption compared to the median number vaccinations missed in a non-disruption year (Fig. 4A). Fewer additional children missed Penta1 and Penta3 vaccinations compared to the median number of vaccinations missed in a non-disruption year (Penta1 2,968 (95% CI 1,621 - 4,271), Penta3 3,683 (95% CI 1,636 – 5,670)) (Fig. 4B-C). The variation in the number of children missed by vaccination in a non-disruption year was much larger than the variation of additionally missed children in 2020 (Fig. S11).

**Figure 4:**
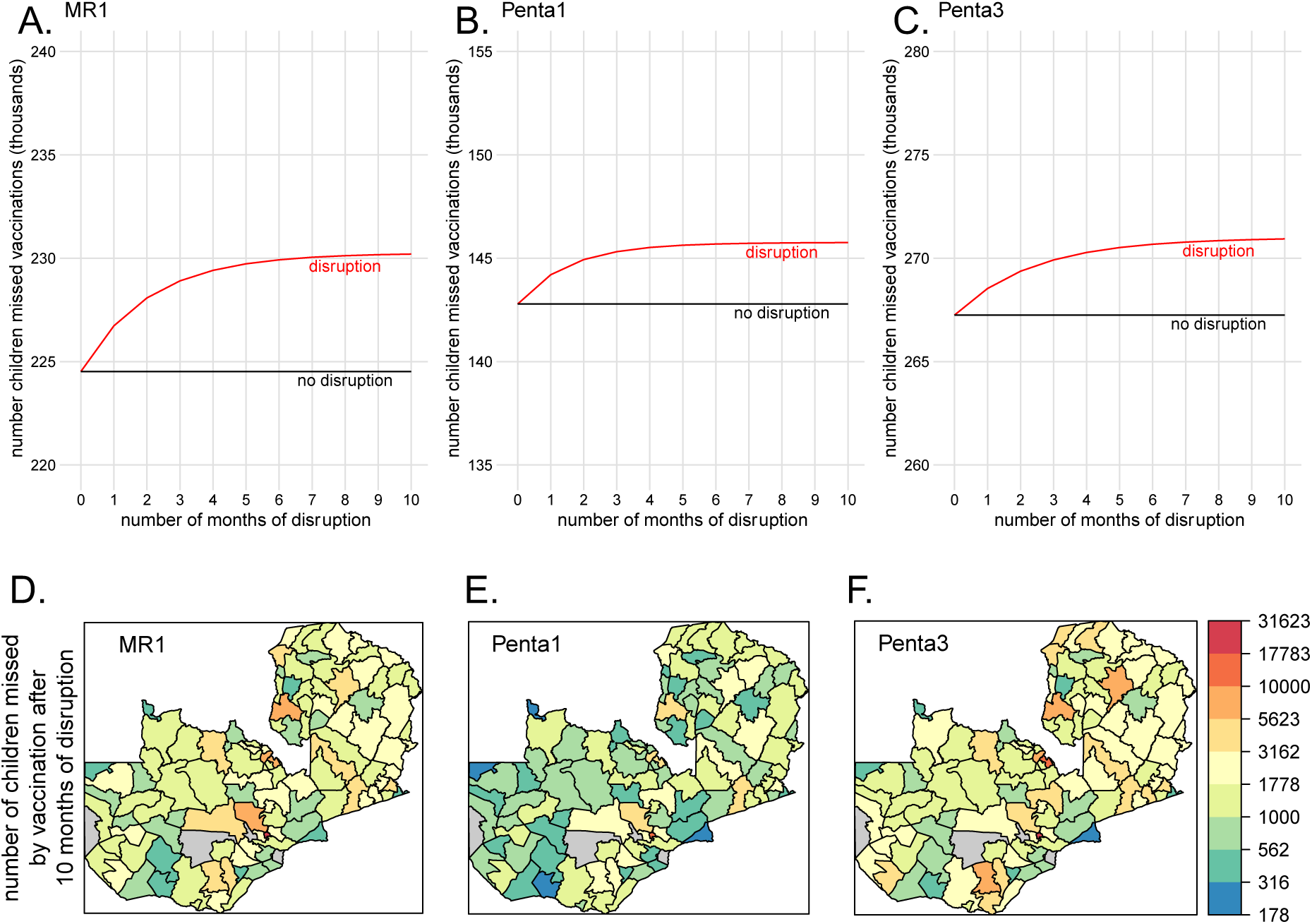
Estimated number of children between the ages of 0 and 36 months missed by vaccination. The national estimated cumulative median number of children missed by vaccination for MR1 (A), Penta1 (B), and Penta3 (C) by month of disruption (red line) across the range of disruption rates (red ribbon) based on the median number of children missed in reference year (black line). Broken down by district (111 / 115 districts) is the median number of children missed by MR1 (D), Penta1 (E), and Penta3 (F) vaccination after 10 months of disruption.

There was large variation in the number of unvaccinated children across districts (Fig. 4D-F). For MR1, Penta1, and Penta 3, Lusaka and Luangwa Districts (both located in Lusaka Province) had the highest and lowest number of children missed in a district, respectively. In Lusaka District, approximately 23,000 children did not receive MR1, 15,500 children did not receive Penta1, and 30,000 children did not receive Penta3. In Luangwa District, approximately 320 children did not receive MR1, 210 children did not receive Penta1, and 300 children did not receive Penta3. There were also a cluster of districts in Copperbelt Province that had a high number of unvaccinated children.

We also evaluated the impact of the COVID-19 pandemic disruptions to routine immunization services on the risk of measles outbreaks for each district over the course of the pandemic prior to the national MR vaccination campaign in November 2020. We found minimal impact on outbreak risk because of pandemic-related disruptions in routine vaccination or delay in fall 2020 MR vaccination campaign. Over the course of 10 months, measles Re increased on average across the districts by 0.05% (range 0.02% - 0.16%) (Fig. 5). There was little to no change in Re during the two-month delay in conduction the MR campaign in all districts (Fig. 5, S5).

**Figure 5:**
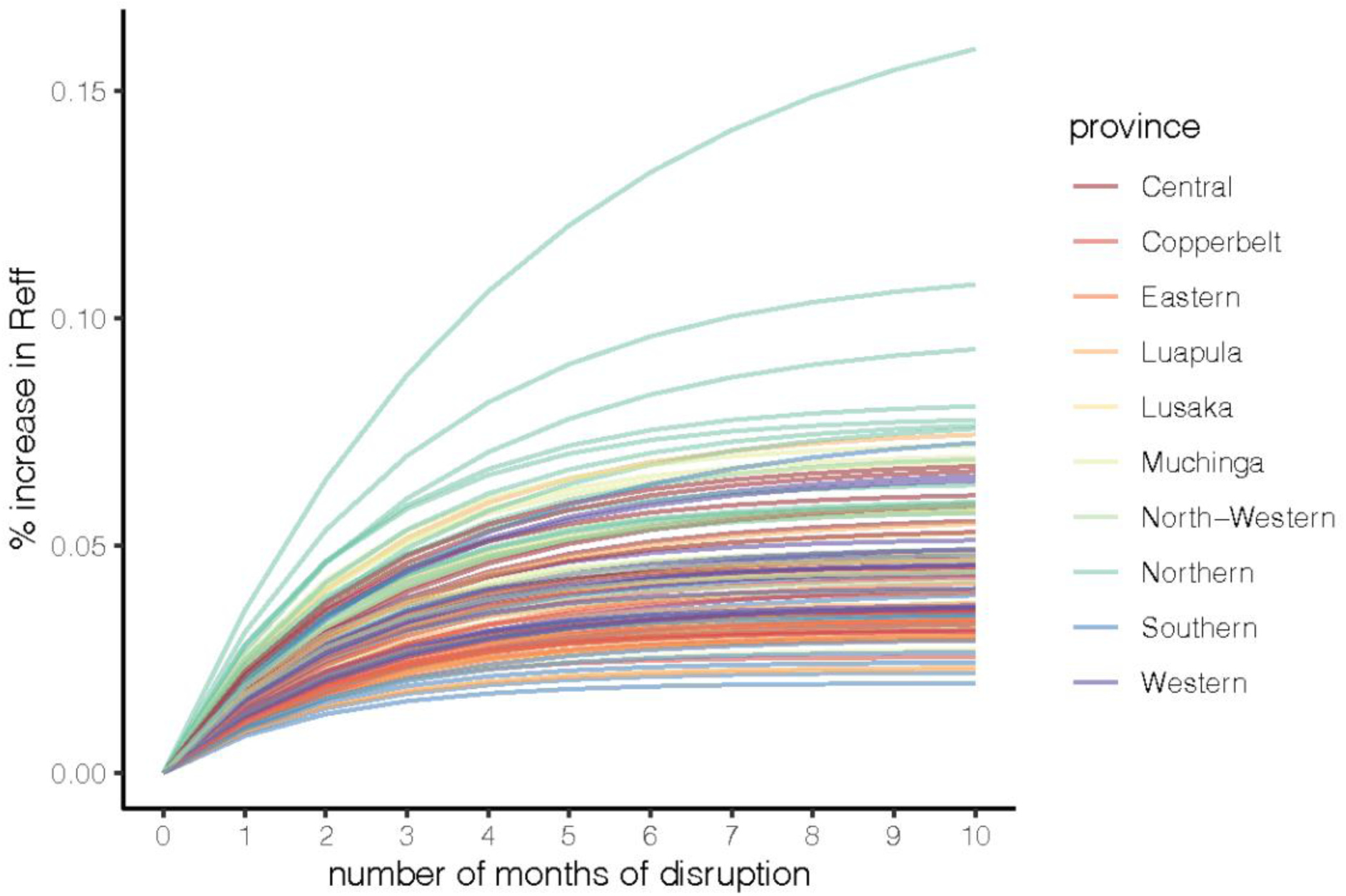
Percent increase in Reff by month of disruption. Each line represents a different district (111 / 115 districts), the color represents the province that each district is located.

While our results focus on the first 10 months of 2020, we also applied our methods to assess potential disruptions in vaccination coverage using updated administrative data between November 2020 and March 2021. A nationwide MR vaccination campaign was conducted in November 2020 targeting all children 9 months through 4 years of age during the biannual child health week resulting in an expected increase in MR doses in November (Fig. S12). However, it is difficult to compare the number of children who received vaccination in November of 2020 to November of 2018 because the target population is inconsistent between the two years (2018 9-23 months, 2020 9-59 months); therefore, the month of November was excluded from the disruptions estimates between November 2020 and March 2021 for MR1. We found that even during a large wave of COVID-19 cases between December 2020 and February 2021 (1), there remained minimal or no disruptions in routine vaccination services (Fig. S12). The mean percent reduction across months was 3.8% (95% CI 2.1, 5.3%), -1.5% (95% CI 0.1, -3%), and -0.7% (95% CI 1.0, -2.0) for MR1, Penta1, and Penta3, respectively.

## Discussion

We performed a detailed analysis of the impact of the COVID-19 pandemic on the routine immunization program in Zambia across time (by month) and space (at administrative level 2). The methodological framework developed in this analysis can be used to evaluate COVID-19 pandemic disruptions in other countries or for other vaccines at a sub-national level. Despite documented impact of the pandemic on vaccination programs in many countries (2), we found minimal disruptions to the ability of the routine immunization program in Zambia to deliver MR1, Penta1, and Penta3 vaccines between January 2020 and March 2021 compared to pre-pandemic levels; a time frame that encompassed two waves of COVID-19 cases in July 2020 and January 2021.

We relied on administrative vaccination data (i.e., the reported number of doses delivered by month and district) and estimated birth cohort size to estimate a mean and range of reductions in the rate routine vaccination. In many countries, including Zambia, health workers at each health facility compile the number of administered vaccine doses manually each month from clinic records including registries, and then send reports to a district health officer. The number of administered doses may be inflated because doses outside the recommended age range are included or deflated because some doses will not be counted. We used WorldPop and Zambia Central Statistical Office demographic projections to estimate the birth cohort (i.e., recommended age range) (11). The estimated size of the birth cohort for each district may be over-or under-estimated depending on the growth rate of the district’s population and in and out migration. As a result, administrative vaccination data is subject to many potential biases and is typically considered less reliable than vaccination coverage surveys (19). To counter this limitation, we relied on survey data (i.e., DHS) to estimate the magnitude of the vaccination rate by district and relied on administrative data to estimate differences in the rate of vaccination between 2018 and 2020. As long as data collection and associated biases in administrative vaccination data and estimated birth cohort are consistent across years, estimated rates of disruptions would be robust to biases within the data.

The administrative vaccination data was not sufficient to evaluate sub-national disruptions to routine vaccinations, so we estimated a national rate of reduction with uncertainty based on district level data. The result is that potential heterogeneities in disruption to routine services over space were not captured. Rather, the variation in the number of unvaccinated children across districts was driven by district-specific size of birth cohorts and baseline vaccination coverage. Importantly, this analysis suggests that variation in the number of unvaccinated children during a non-disruption year was much larger than the variation in unvaccinated children during the COVID-19 pandemic disruptions in 2020. This finding further highlights the minimal impact of the COVID-19 pandemic on routine immunization services in Zambia, as well as the substantial between-district variation in the lifetime probability of vaccination with MR1, Penta1, and Penta3.

Continued routine and catch-up immunization services during the pandemic have shown to be a net benefit in modelling studies (3). Regardless, there remain concerns about the potential for SARS-CoV-2 transmission during routine or campaign vaccination activities as a result of interactions with healthcare workers or other individuals seeking services. Zambia’s Child Health Weeks conducted in June and November of 2020 took place despite the pandemic. These vaccination activities in Zambia included COVID-19 precautionary protocols including use of PPE and minimum distance requirements for individuals seeking care. The purpose of Zambia’s Child Health Week, held biannually, is to reach eligible children who had not yet received their routine vaccines. This purpose was critically fulfilled during the pandemic when the June 2020 Child Health Week resulted in a greater increase in the number of vaccinated children from the baseline 2018 year for MR1 and Penta3. The MR vaccination campaign that was delayed two months was instituted during Zambia’s second yearly Child Health Week in November 2020.

This vaccination campaign / Child Health Week was charged with not just capturing children who had not revied their routine services, but to vaccinate all individuals in the campaigns’ target age range (9 months-4 years). The impact of the MR campaign on measles and rubella susceptibility will depend on the correlation in the probability of receiving routine vaccination versus campaign vaccination. Initial findings suggest that the November 2020 vaccination campaign reached more children than the November 2018 Child Health Week (Fig. S12); however, it is unknown if these children were previously unvaccinated. The approach used in Zambia demonstrates the benefits of continuing with routine immunization services during the pandemic and using catch-up vaccination activities to vaccinate those children who may have missed due to COVID-19 pandemic disruptions.

We focused on the potential of the COVID-19 pandemic to disrupt vaccination programs negatively. However, there are other potential ways the pandemic can impact the burden of vaccine-preventable diseases. For example, non-pharmaceutical interventions, such as movement restrictions (either personal or state enforced), can reduce transmission of directly transmitted infectious pathogens. Minimal disruption of routine vaccination programs coupled with decreased movement and transmission of vaccine-preventable diseases can inadvertently lead to local eliminations with minimal additional risk of resurgence. However, decreased circulating viruses can also create a short-term illusion of control without considering the potential risk of increasing susceptible populations. As a result, and regardless of the pandemic, it is important that vaccination coverage continues to improve in Zambia for all birth cohorts. Nationally, vaccination coverage of MR1 is high (96% in 2020) but MR2 coverage has plateaued in recent years around 65% (66% in 2020) (20). While vaccine effectiveness is fairly high for one dose of MR (84% measles and 97% rubella), a second dose of MR with 97% vaccine effectiveness is critical to control measles.

Ongoing collaborations and established research programs on measles and rubella in Zambia allowed a timely assessment of the impact of COVID-19 disruptions on routine immunization services and the impact of delaying a MR vaccination campaign for two months. For example, rich measles serological data collected in 2016 from a national serosurvey was used to set a baseline R effective for assessing changes in measles outbreak risk over months of disruption (15). We identified minimal increases in risk of a measles outbreak due to postponing the MR vaccination campaign by two months. The campaign was indeed delayed with no outbreaks reported over the course of those two months. However, it is important to note that we focused on the district level change in outbreak risk which could be averaging across potential within-district heterogeneities in susceptibility (21). It is also worth noting that ideally we would have been able to report districts’ predicted risk of a measles outbreak during the pandemic, rather than simply the change in risk. However, the hierarchical seroprevalence model was not suitable to make such predictions. The reliance on the model’s district-specific random intercept would have required a strong assumption that the underlying district-specific impact on seroprevalence is constant from 2016 to 2020. A key area of future work is to build models that can reliably extrapolate seroprevalence to other years from rich population-based cross-sectional serological data. As suggested by this analysis, this may require individual level data on mechanisms of seroconversion (i.e., history of vaccination or measles infection) linked to the serum samples. Another avenue is to incorporate serological data (potentially from smaller more spatially targeted serosurveys or from residual specimens) from the years following the cross-sectional survey into a time-specific seroprevalence model.

Since this analysis was completed, there has been an additional wave of COVID-19 cases that peaked in July 2021. The implications of the pandemic on Zambia’s childhood vaccination program are not fully realized. Program managers and policy makers must take advantage of every opportunity to provide catch-up vaccination and re-engage families with the health care system. Further analysis is needed to evaluate the ongoing disruptions and understand potential subdistrict variations in the impact of the pandemic on childhood vaccinations.

## Data Availability

Data produced in the present study can be available upon reasonable request to the authors

