## Supplement for "An evaluation of the early impact of the COVID-19 pandemic on Zambia’s routine immunization program"

#### ***Appendix S1.*** *Estimating baseline routine vaccination coverage for MR1, Penta1, and Penta 3*

We relied on data on routine vaccination coverage from Zambia's 2018 cross-sectional Demographic and Health Survey (DHS) made publicly available by ICF International to estimate the lifetime probability and monthly rate of receiving routine vaccination for each district in Zambia. The national DHS survey has one record for each interviewed woman's child aged 0 to 36 months at the time of the survey (N=5,670), and is linked to a database of GPS coordinates (latitude and longitude) of respondents' home locations. The coordinates are aggregated on the scale of clusters containing approximately twenty households, and randomly jittered up to 2 kilometers in urban areas and up to 5 kilometers in rural areas to protect respondent confidentiality (however, the jittered coordinates remain within their respective administrative districts). Children included in the 2018 DHS survey were too young to have been eligible for vaccination in the most recent measles supplementary immunization activity (SIA) campaign conducted in 2016, so we focus here on routine vaccination only.

For each child, we extracted from the DHS: age at the time of survey, whether the child had ever received a MR1, Penta1, and Penta3 vaccine (each based on either vaccination card or report of parent/guardian), and age at the time of the vaccine (if a vaccination card was available). Data is available from 111 of the 115 designated districts in 2018. For MR1, Penta1, and Penta3, children under 8, 0.5, and 2.5 months of age at the time of DHS survey, respectively, were excluded from the analysis; they were considered not to be "at risk" for vaccination since recommended MR1 administration is at 9 months, 6 weeks, and 14 weeks age, respectively, in Zambia. We modified a survival analysis approach developed in [1,2] to estimate parameters associated with vaccination coverage. A child with a vaccination card was considered to be uncensored at the time of the vaccination (we assume that vaccinations recorded on cards only represent routine vaccination). When a parent reported that their child had received vaccination but a vaccination card was unavailable, the child was considered to be left censored at the time of DHS survey; a child reported as unvaccinated was considered to be right censored at the time of DHS survey.

For an individual $j$ at district $i$, the probability that this individual has been vaccinated depends on three classifications of individuals: uncensored ($u_{ij} = 1$), left censored ($l_{ij} = 1$), or right censored ($r_{ij} = 1$):

$$f(v_{ij}; \theta_i) = Pr(T = v_{ij}) = \lambda_i \cdot e^{-\lambda_i \cdot (v_{ij} - m)} \quad \text{(Eq. 1)}$$

$$F(t_{ij}; \theta_i) = Pr(T \leq t_{ij}) = p_i \left(1 - e^{-\lambda_i \cdot (t_{ij} - m)}\right) \quad \text{(Eq. 2)}$$

$$S(t_{ij}; \theta_i) = Pr(T > t_{ij}) = 1 - F(t_{ij}; \theta_i) \quad \text{(Eq. 3)}$$

$T$ is a continuous random variable representing the age in months at receipt of vaccine; $v_{ij}$ is the age in months at the receipt of vaccine, only available for uncensored individuals ($u_{ij} = 1$); $t_{ij}$ is the age in months of an individual at the time of the DHS survey; $p_i$ is the location-specific lifetime probability of being vaccinated through routine vaccination (e.g., the saturation parameter); $\lambda_i$ is the location-specific rate of receiving the vaccine through routine vaccination after *m* months of age; *m* is vaccine-specific and is 8 for MR1, 0.5 for Penta1, and 2.5 for Penta3. The probability of routine vaccination reaches $p_i$ more slowly (e.g., at an older age) for lower values of $\lambda_i$ than it does for higher values (Fig. S1).

The parameters of interest, transformed for stability, are: $\theta_{1,i} = logit(p_i)$, and $\theta_{2,i} = log\ (\lambda_i)$. The likelihood of the parameters given the observed age and vaccination status of an individual is:

$$L(\theta_i; t_{ij}, v_{ij}, u_{ij}, r_{ij}, l_{ij}) = f(v_{ij}; \theta_i)^{u_{ij}} \cdot F(t_{ij}; \theta_i)^{u_{ij} + l_{ij}} \cdot S(t_{ij}; \theta_i)^{r_{ij}} \qquad \text{(Eq. 4)}$$

To estimate $\theta_{x,i}$ we assumed that it is multivariate normally distributed with a location-specific mean of $\mu_{x.i}$ (Eq. 5) and a conditional autoregressive model (CAR) specification for the spatial random effects, parametrized by the precision matrix $1/\sigma_x^2$:

$$\theta_{x,i} \sim Normal(\mu_{x,i}, \sigma_x^2) \qquad \text{(Eq. 5)}$$
$$1/\sigma_x^2 = \tau_x(D - \alpha_x W) \qquad \text{(Eq. 6)}$$

Where $1/\sigma_x^2$ is the precision matrix, $\tau_x$is a precision parameter, $\alpha_x$controls the spatial dependence ($\alpha$=0 implies spatial independence, and $\alpha$=1 collapses to an intrinsic conditional autoregressive model), $D$ is an *i* by *i* diagonal matrix with diagonal elements encoding the number of adjacent neighbors that each district has, and $W$ is a binary adjacency matrix. Because the distribution of the conditional autoregressive model is multivariate normal, we only include a spatial random effect in these models.[3]

### ***Appendix S2.*** *Estimating R effective*

To estimate outbreak risk we calculated the measles effective reproduction number ($R_e$) for each month and district using next generation methods taking into account age-specific susceptibility and mixing patterns.[4] The next generation matrix **W**, with elements $w_{ij}$, represents the expected number of individuals in the *i*th age class that are infected by an infectious individual in the *j*th age class upon introduction of measles virus into a totally susceptible population. The basic reproduction number, or the average number of individuals infected by a typical infected individual, is given by $R_0 = p(A)$ where $p(A)$ represents the dominant eigenvalue of matrix A. We initially calculate **W** by scaling age contacts extracted per [5] to a conservatively assumed basic reproduction number of measles of 12 (although estimates vary widely [6]), such that $p(\boldsymbol{W}) = 12$. To estimate $R_e$ for each district *d* and month of disruption *m* we reevaluate the dominant eigenvalue of **W** after multiplied by the proportion of susceptible population,

$$R_{e_{dm}} = \rho \begin{bmatrix} w_{11}S_{1_{dm}} & \cdots & w_{1z}S_{1_{dm}} \\ \cdots & \cdots & \cdots \\ w_{z1}S_{z_{dm}} & \cdots & w_{zz}S_{z_{dm}} \end{bmatrix} \qquad \text{(Eq. 7)}$$

where, $S_{k_{dm}} = S_k(dm) / N_k(d)$ is the proportion of susceptible population in age-group *k* specific to district *d* and month of disruption *m*. The size of the population in age-group *k* specific to district *d* ($N_k(d)$) was assumed constant throughout the year 2020 using Zambia Central Statistical Office estimates.

### ***Appendix S3.*** *Estimating measles susceptibility in 4 to 49 years olds using a seroprevalence model*

To estimate district-specific measles seroprevalence, we applied an approach developed in [7] where hierarchical spatial models were fit to data on individual measles seropositivity from an analysis of residual samples from a national HIV serosurvey (Zambia Population-Based HIV Impact Assessment) in Zambia in 2016.[7] The nested serosurvey consisted of 9,852 blood samples collected from individuals one month to 49 years old. District-specific random effects were included in the hierarchical model based on a conditional autoregressive (CAR) specification, meaning that estimated seroprevalence at any given district was conditional on the estimated seroprevalence of neighboring districts. The models assumed a binomial probability distribution for seropositivity and a log odds link. Informative epidemiological and demographic model covariates included: individual level HIV status and age, and district and age specific measles vaccine dose 1 coverage and exposure to local measles outbreaks. There was no information about individuals' history of measles vaccination or measles infection from the nested serosurvey; this meant we had to rely on population level (i.e., district and age specific) covariates associated with history of measles vaccination and measles infection. Measles vaccine dose 1 coverage was estimated from Zambia DHS 2013 data. We defined exposure to an outbreak as any individual alive and living in a district with two or more measles-specific IgM positive cases reported within a year. Measles case data was collected by Zambia EPI program and was available since 2012. We also included interactions between HIV serostatus and age, as well as HIV serostatus and squared age. See [7] for additional model details. To project measles seroprevalence in year 2020 (for use in this analysis), data specific to modeled covariates from 2020 were combined with posterior estimates of model parameters.

We conducted leave-one-out cross validation analyses to evaluate the performance of the model to predict district and age-specific seroprevalence. In these analyses we left out each district or age in years, retrained the model with the smaller dataset, and then used the new posterior estimates of the parameter values to predict seroprevalence for the district or age originally left out (Fig. S3-S4). The model does well to predict seroprevalence for missing ages, but not as well to predict seroprevalence for missing districts. This finding is expected, given our reliance on the model district-specific intercept (gamma parameter) that captures variation not explained by our demographic and epidemiologic covariates. We do not extrapolate our model to estimate seroprevalence for any new districts. Our extrapolations for 2020 estimates of seroprevalence rely on the assumption that the underlying district-specific impact on seroprevalence (i.e., district-specific intercept) is constant from 2016 to 2020. As a result, we only evaluate the change in R effective due to routine vaccination disruptions in each district and do not evaluate the magnitude of R effective itself. We also present sensitivity analyses of the change in R effective to assumptions about population susceptibility in 2020 (Fig. S5).

We additionally compared estimated seroprevalence in 2020 for ages 9 to 36 months old to estimated immunity derived from vaccination (i.e., age and district-specific vaccination from the DHS estimates above taking into account vaccine effectiveness). We found generally good agreement between the two estimates; 95% credible intervals overlapped in at least half of the ages (>=14 of 28 age groups) in 95% of districts (106 of 111) and 95% credible interval of immunity (via DHS) overlapped with the median estimated seroprevalence in at least half of the ages (>=14 of 28 age groups) in 47% of districts (52 of 111) (Fig. S6). Given that estimates of immunity from coverage data only includes immunity due to vaccination, we would expect there to be good agreement in districts where there is little natural infection (i.e., Zimba district, Fig. S6B) and lower than seroprevalence in districts where natural infection may still contribute to immunity (i.e., Chitambo district, Fig. S6B). However, in the majority of districts (including Lusaka, Fig. S6B), estimates of immunity from DHS data are higher than seroprevalence. This may suggest that i) DHS data is inherently biased to capture children who access healthcare and vaccination, ii) immunity levels waned very quickly for vaccinated children or the EIA kit has a lower sensitivity than documented by the manufacturer, or iii) model fitting issues causes biases in one or both estimates. Although we cannot totally rule out that potential that vaccine effectiveness is much lower in Zambia during this time for than other populations or that the sampling units for each survey differ across districts and represent drastically different populations within a district.

The final step to estimate district-specific seroprevalence is to weight the seroprevalence estimates by district-specific population characteristics. We created a new dataset with all possible covariate groupings. For example, one possible covariate grouping is district Chadiza, province Eastern, HIV negative, 7 years old, and with no exposure to a local outbreak. We estimated the probability of seropositivity for each covariate grouping across 2500 samples from parameter posterior distribution sets, taking into account both uncertainty of the mean parameter values and uncertainty in the sampling process. We then weighted the probability of seropositivity by each covariate grouping and sampled parameter set per age and district by the proportion of individuals in that covariate grouping to get 2500 estimates of seroprevalence for each age and district.

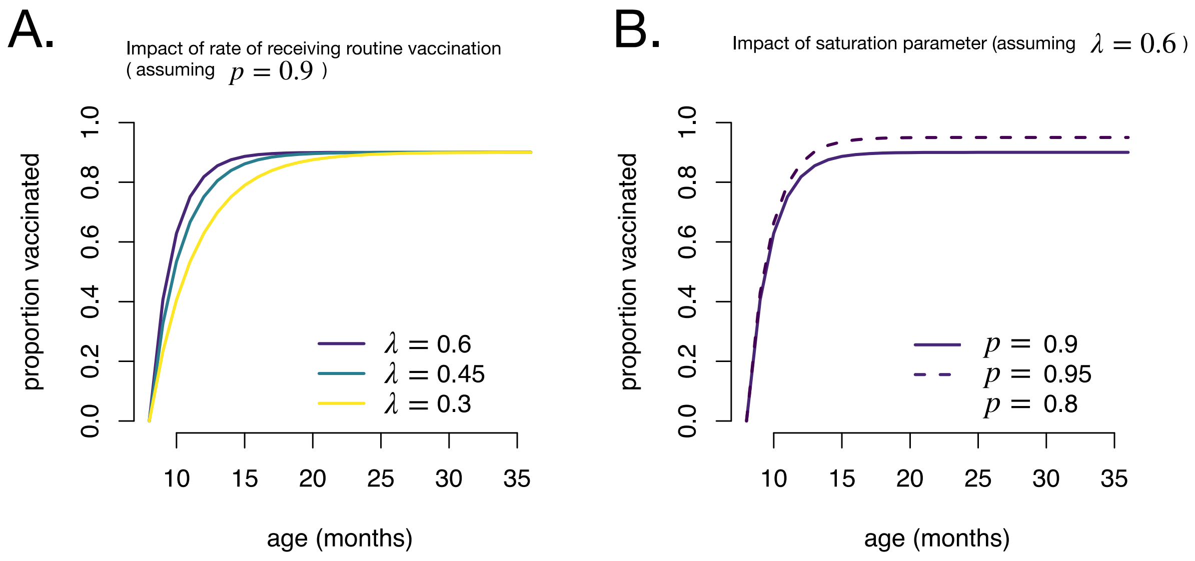


**Figure S1:** Impact of estimated parameters $\lambda$ (**A**) and $p$ (**B**) on the proportion vaccinated over age.

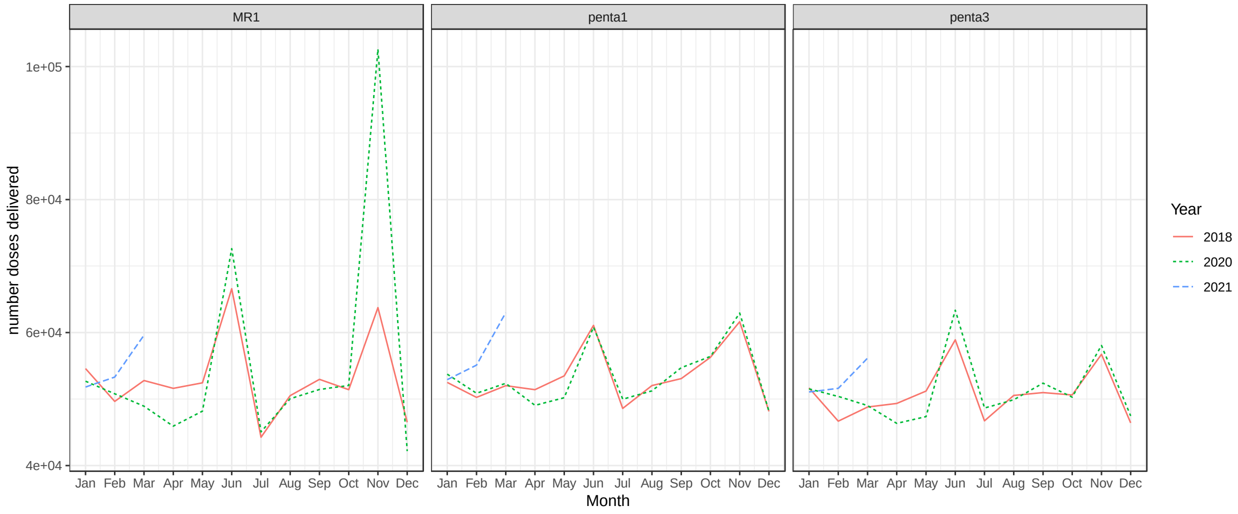


**Figure S2:** Raw adminstrative vaccination data. The number of MR1, Penta1, and Penta3 vaccine doses adminstered from January to December. Each line represents a different year of data. Child health weeks are conducted in June and November of each year, hence the spikes in vaccine doses in these months.

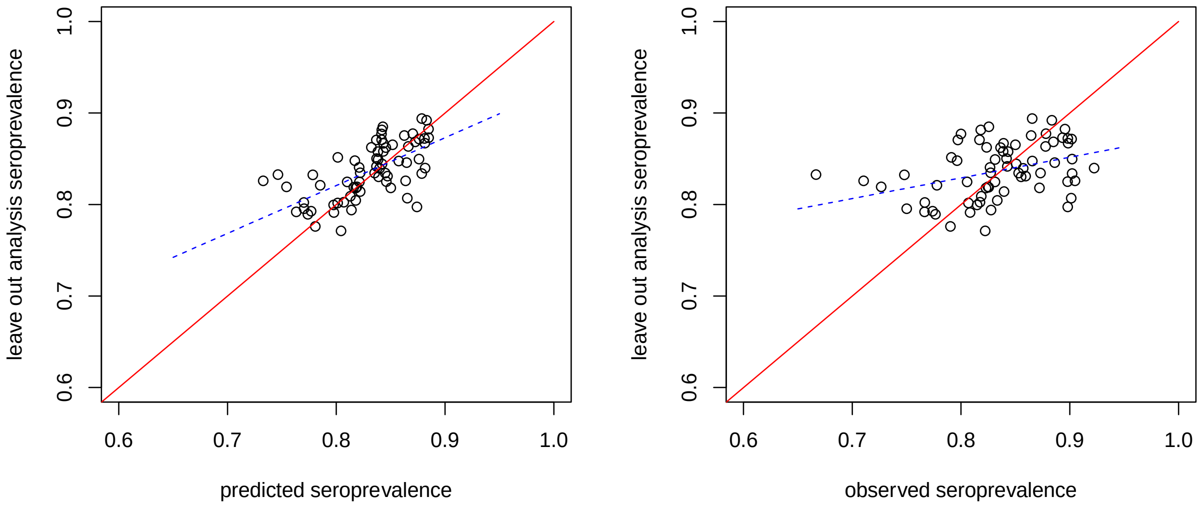


**Figure S3.** Results of leave out district analysis. Each point represents a different district. Left figure displays the estimated mean seroprevalence for a predicted districts left out of the analysis by the expected seroprevalence given the district was included in the analysis. Right figure displays the estimated mean seroprevalence for a predicted districts left-one-out of the analysis by the observed seroprevalence for the respective district. Dashed blue line is fit line and red solid line represents perfect agreement.

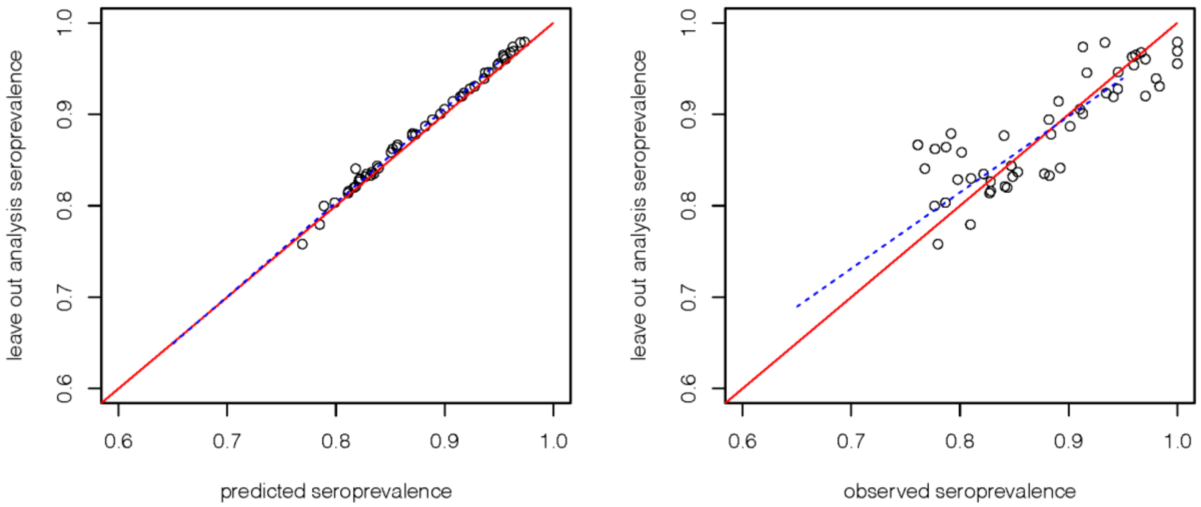


**Figure S4.** Results of leave out age analysis. Each point represents a different age in years. Left figure displays the estimated mean seroprevalence for a predicted ages left out of the analysis (y-axis) by the expected seroprevalence given the age was included in the analysis (x-axis). Right figure displays the estimated mean seroprevalence for a predicted ages left-one-out of the analysis (y-axis) by the observed seroprevalence for the respective age (x-axis). Dashed blue line is fit line and red solid line represents perfect agreement.

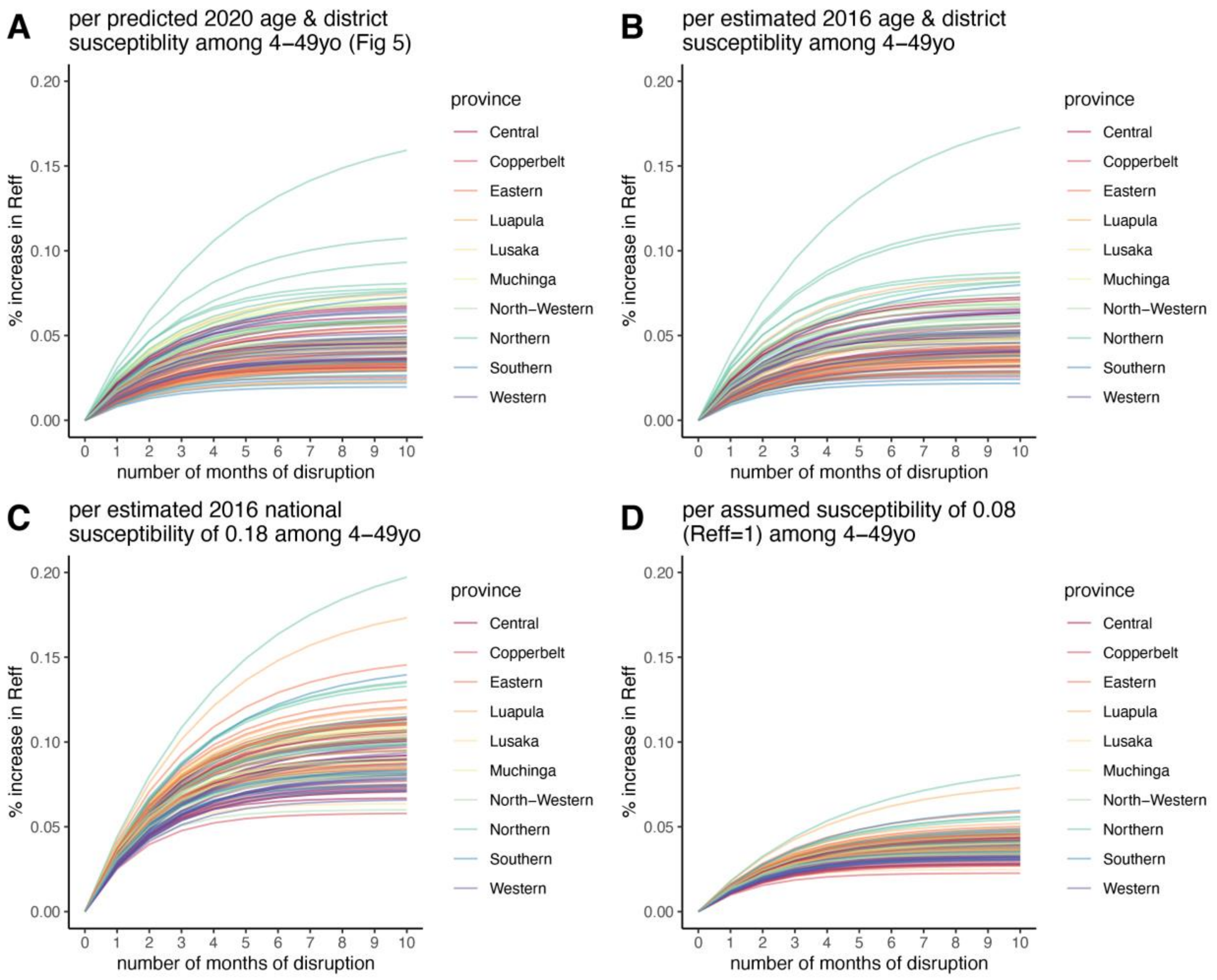


**Figure S5**. Sensitivity analysis of percent increase in Reff per month of disruption based on different starting estimates of proportion susceptible across ages 4 to 49 years old. **A)** same as main text Figure 5 is based on predicted 2020 age- and district-susceptibility among 4 to 49 years old. **B)** based on 2016 age- and district-specific susceptibility among 4 to 49 years old. **C)** based on estimated 2016 national susceptibility of 0.18 assumed for all ages 4 to 29 years old. **D)** based on assumed susceptibility of 0.08 (equivalent to Reff=1) assumed for all ages 4 to 49 years old.

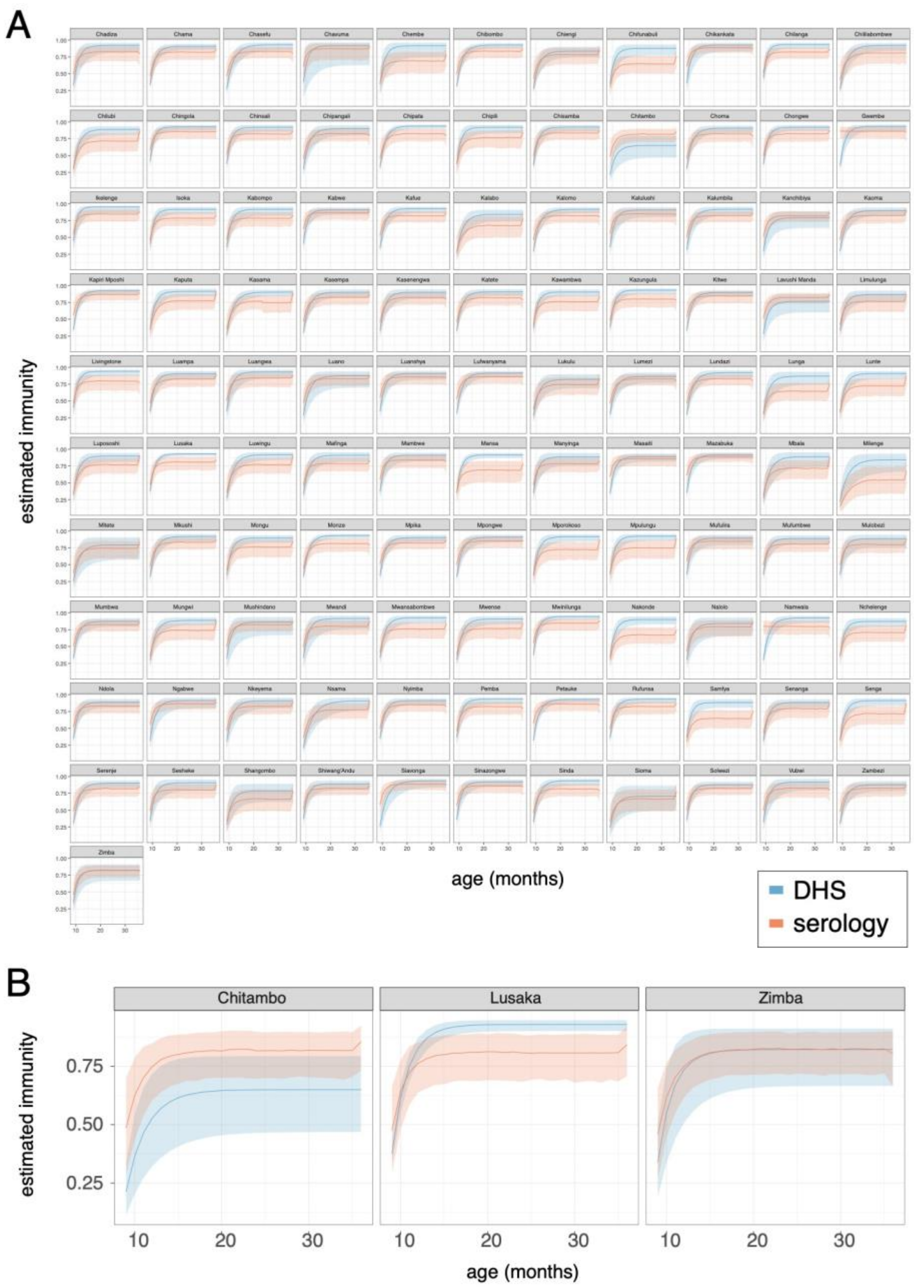


**Figure S6**. Comparison of estimated immunity by age (9-36 months) using seroprevalence data and demographic and heath survey data. The described above ‘measles seroprevalence model’ is a hierarchical spatial models fit to individual measles seropositivity in 2016 that was used to predict age and district specific seroprevalence in 2020; the median and 95% credible intervals are displayed in orange line and transparent orange ribbon, respectively. The section above titled ‘baseline routine vaccination’ describes a spatial model fit to 2018 DHS vaccination data that was used to estimate age and district specific immunity (taking into account vaccination effectiveness and minimal disruptions); the mean and 95% credible intervals are displayed in blue line and transparent blue ribbon, respectively. **A)** 111 districts, **B)** 3 districts representing serology estimated immunity higher than DHS coverage estimated immunity, (Chitambo), serology estimated immunity lower than DHS coverage estimated immunity (Lusaka), and good agreement (Zimba).

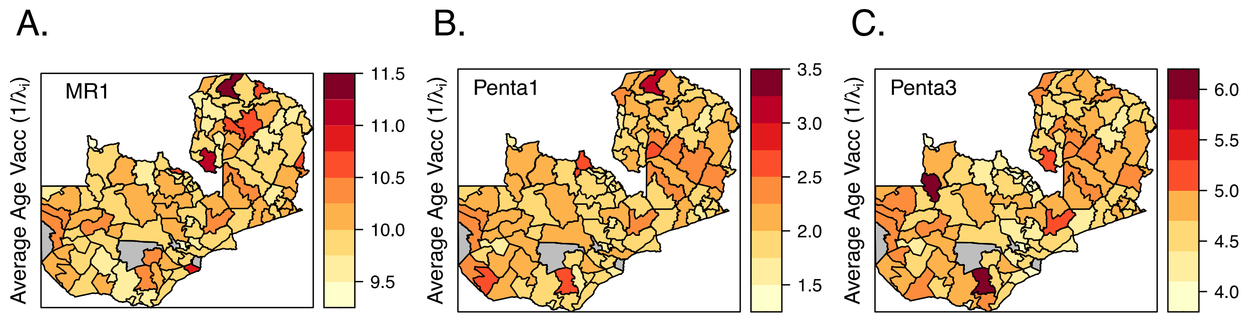


**Figure S7**: District-level estimates of the average age of vaccination among those that receive vaccination for MR1 (A), Penta1 (B), and Penta3 (C). Average age is estimated as the inverse of the median rate of receiving routine vaccination. There are no parameter estimates for 4/115 districts colored in grey due to the lack of any DHS sampling clusters in these districts.

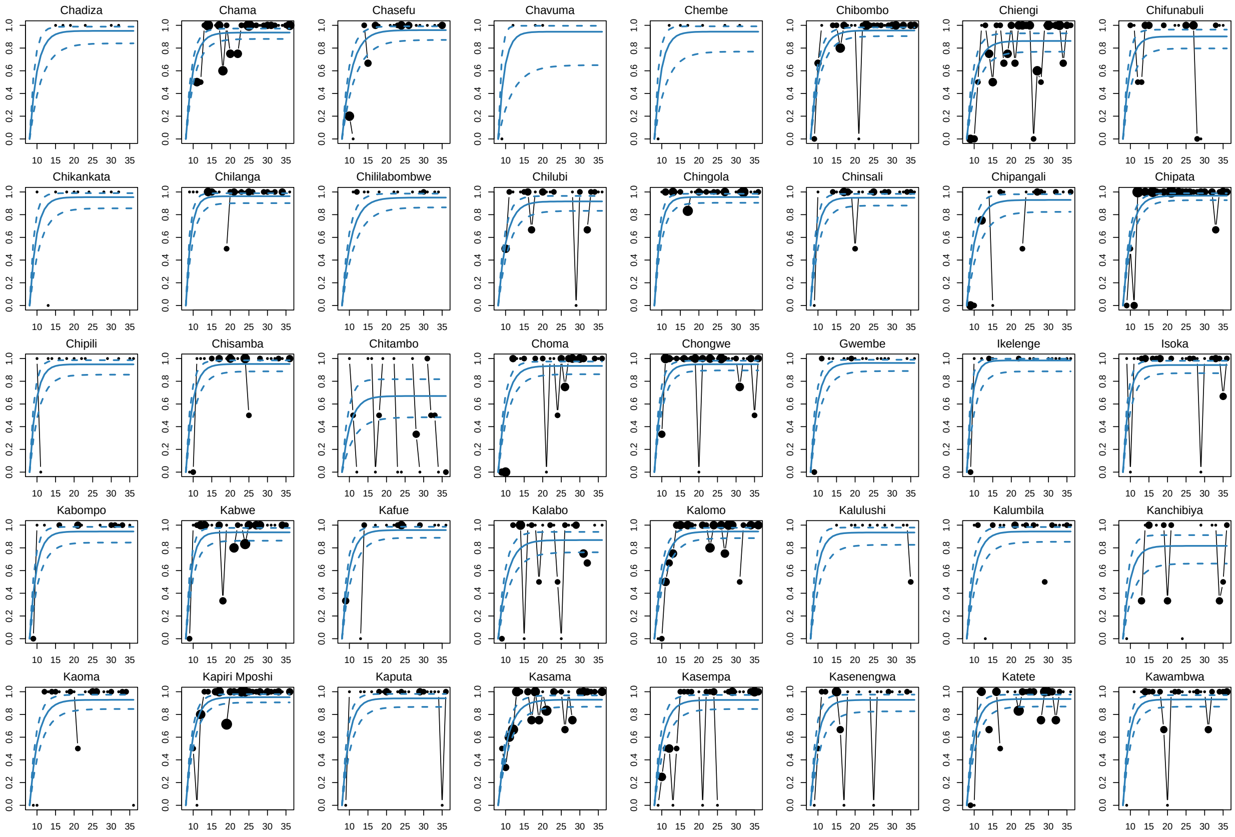
Chadiza
Chama
Chasefu
Chavuma
Chembe
Chibombo
Chiengi
Chifunabuli
Chikankata
Chilanga
Chililabombwe
Chilubi
Chingola
Chinsali
Chipangali
Chipata
Chipili
Chisamba
Chitambo
Choma
Chongwe
Gwembe
Ikelenge
Isoka
Kabompo
Kabwe
Kafue
Kalabo
Kalomo
Kalulushi
Kalumbila
Kanchibiya
Kaoma
Kapiri Mposhi
Kaputa
Kasama
Kasempa
Kasenengwa
Katete
Kawambwa

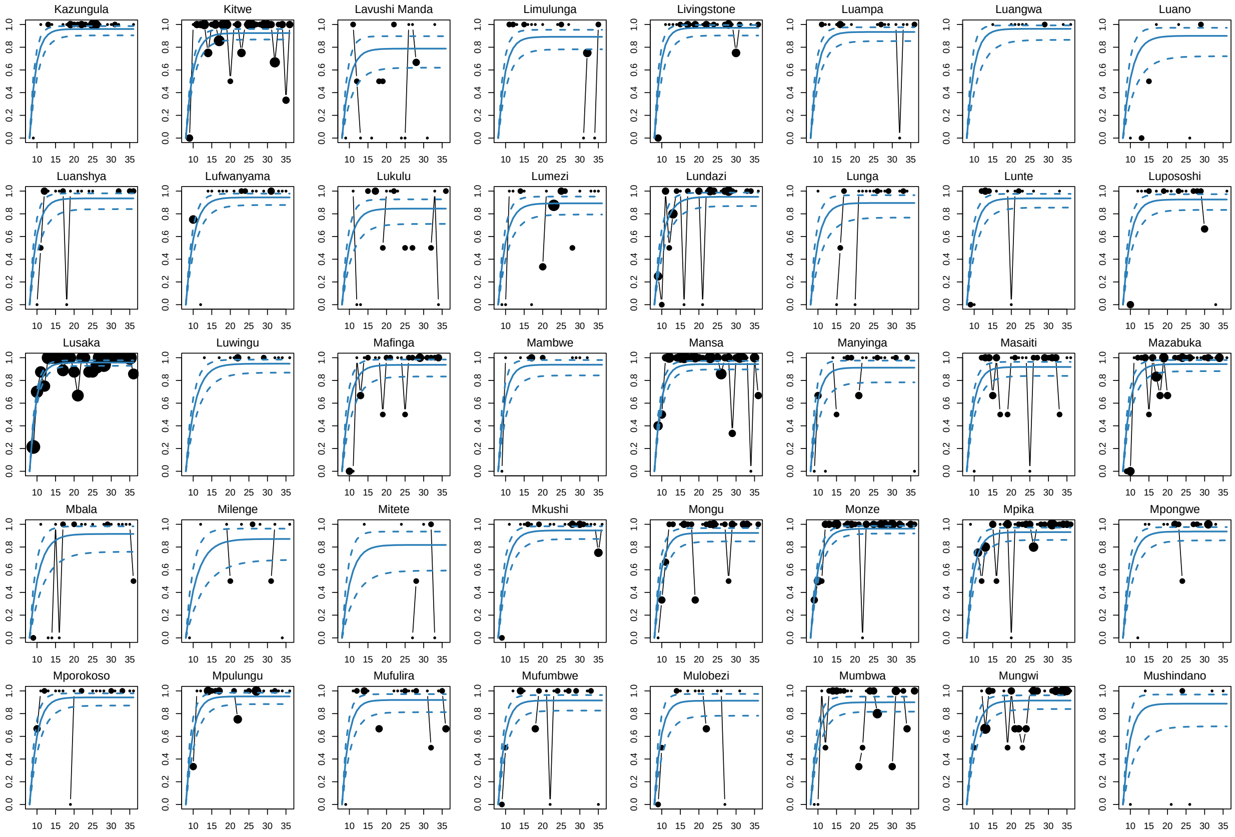
Kazungula
Kitwe
Lavushi Manda
Limulunga
Livingstone
Luampa
Luangwa
Luano
Luanshya
Lufwanyama
Lukulu
Lumezi
Lundazi
Lunga
Lunte
Lupososhi
Lusaka
Luwingu
Mafinga
Mambwe
Mansa
Manyinga
Masaiti
Mazabuka
Mbala
Milenge
Mitete
Mkushi
Mongu
Monze
Mpika
Mpongwe
Mporokoso
Mpulungu
Mufulira
Mufumbwe
Mulobezi
Mumbwa
Mungwi
Mushindano

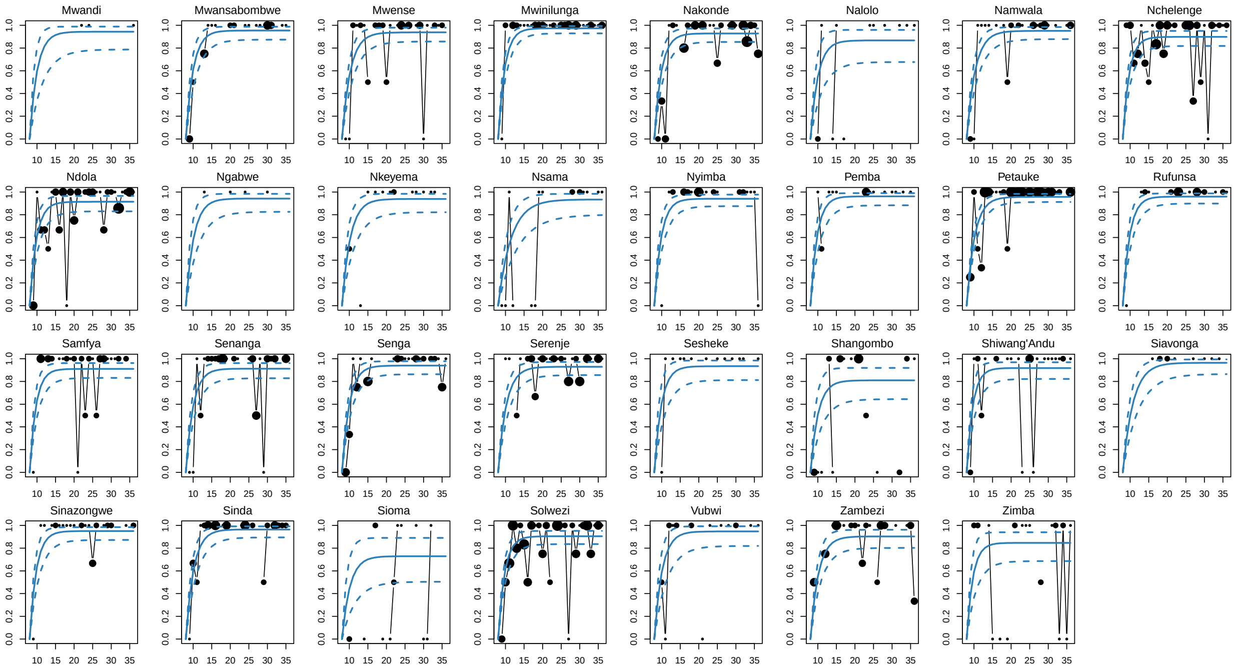


**Figure S8**: District-level MR1 baseline routine proportion vaccinated (y-axis) over age in months (x-axis). Each plot represents one of the 111 modeled districts. The black points represent the data where the size of the point is proportional to the number of observations for each age in months. The solid and dashed lines represent the model fit median and 95% credible intervals.

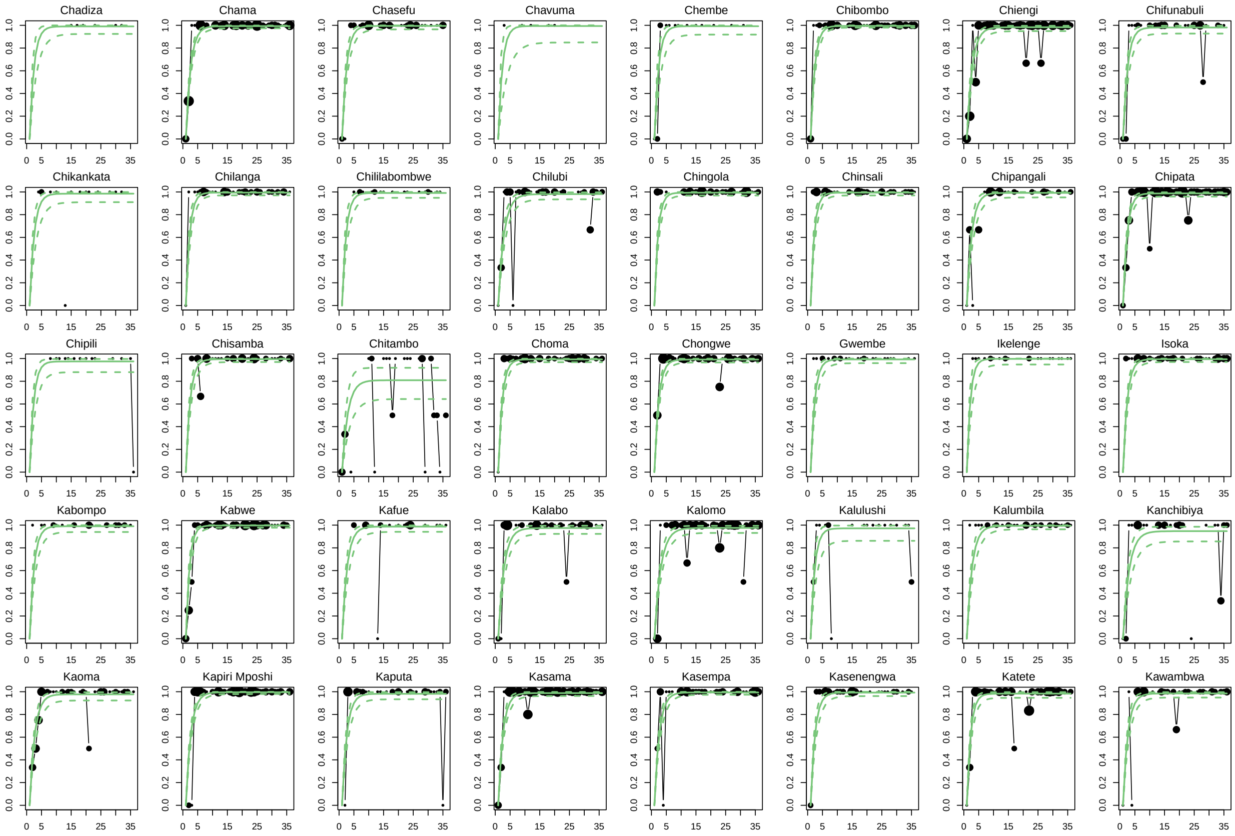
Chadiza
Chama
Chasefu
Chavuma
Chembe
Chibombo
Chiengi
Chifunabuli
Chikankata
Chilanga
Chililabombwe
Chilubi
Chingola
Chinsali
Chipangali
Chipata
Chipili
Chisamba
Chitambo
Choma
Chongwe
Gwembe
Ikelenge
Isoka
Kabompo
Kabwe
Kafue
Kalabo
Kalomo
Kalulushi
Kalumbila
Kanchibiya
Kaoma
Kapiri Mposhi
Kaputa
Kasama
Kasempa
Kasenengwa
Katete
Kawambwa

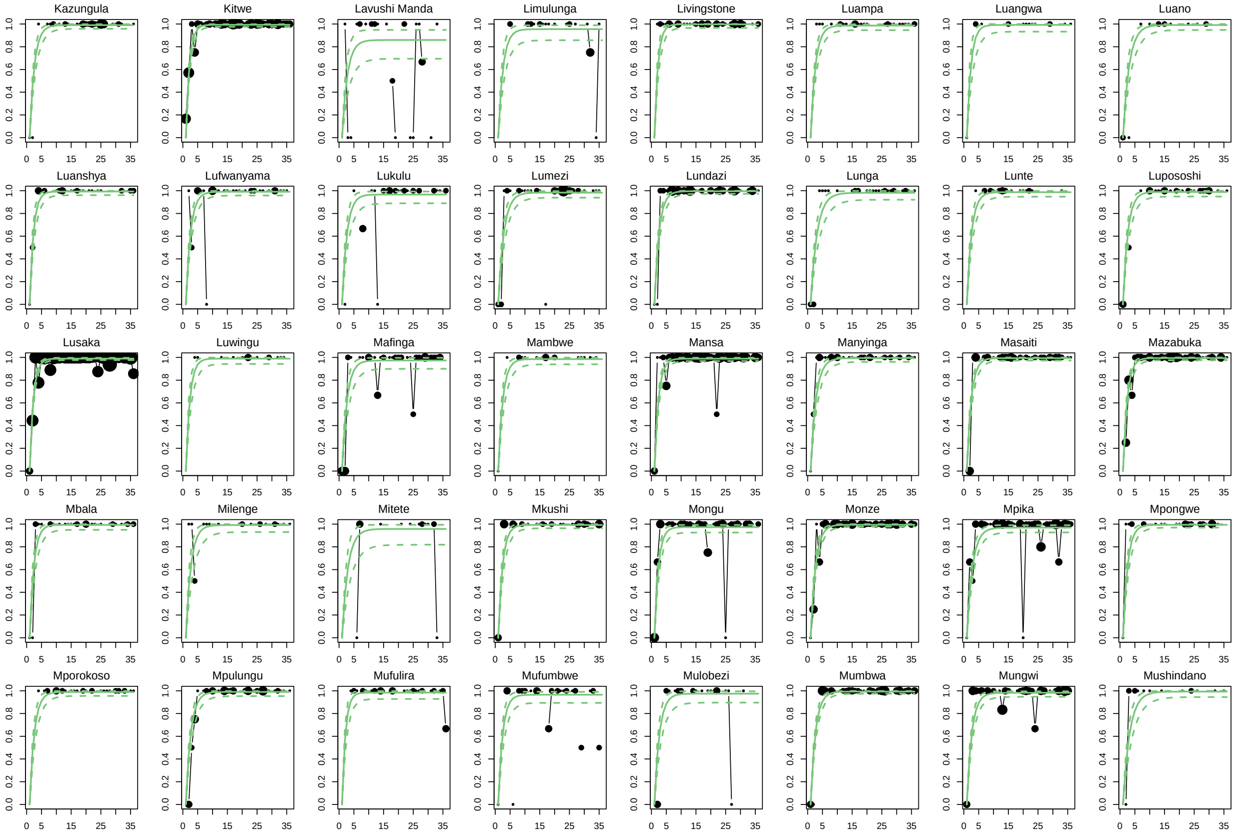
Kazungula
Kitwe
Lavushi Manda
Limulunga
Livingstone
Luampa
Luangwa
Luano
Luanshya
Lufwanyama
Lukulu
Lumezi
Lundazi
Lunga
Lunte
Lupososhi
Lusaka
Luwingu
Mafinga
Mambwe
Mansa
Manyinga
Masaiti
Mazabuka
Mbala
Milenge
Mitete
Mkushi
Mongu
Monze
Mpika
Mpongwe
Mporokoso
Mpulungu
Mufulira
Mufumbwe
Mulobezi
Mumbwa
Mungwi
Mushindano

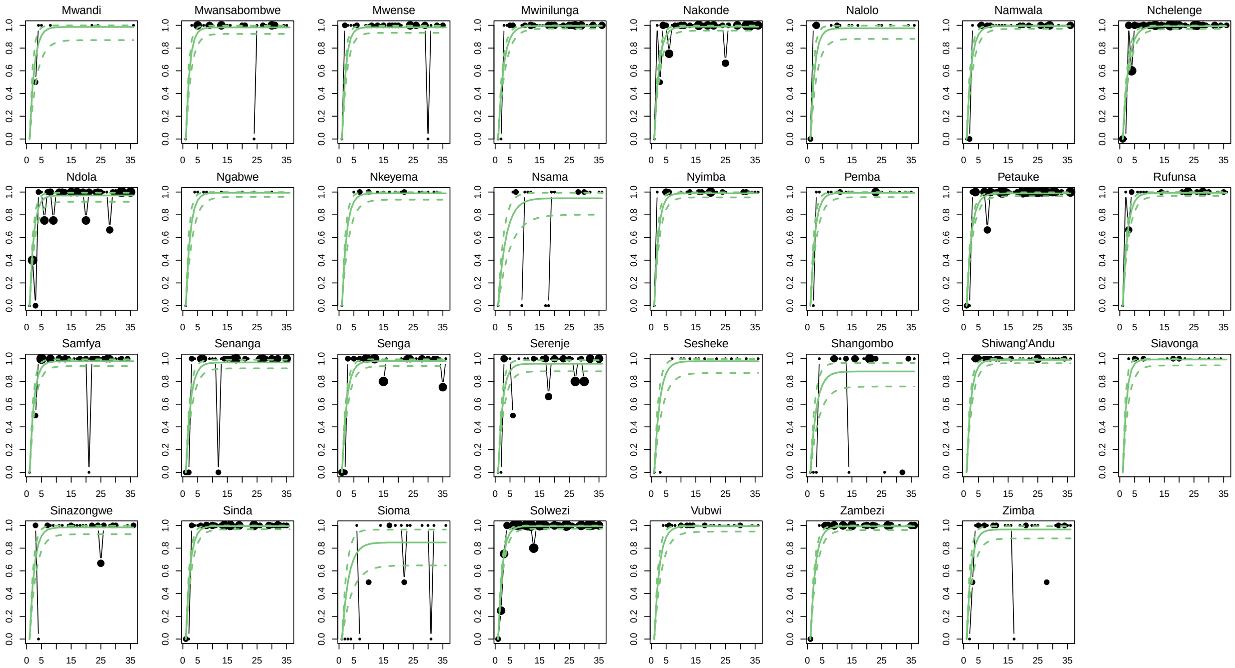


**Figure S9**: District-level Penta1 baseline routine proportion vaccinated (y-axis) over age in months (x-axis). Each plot represents one of the 111 modeled districts. The black points represent the data where the size of the point is proportional to the number of observations for each age in months. The solid and dashed lines represent the model fit median and 95% credible intervals.

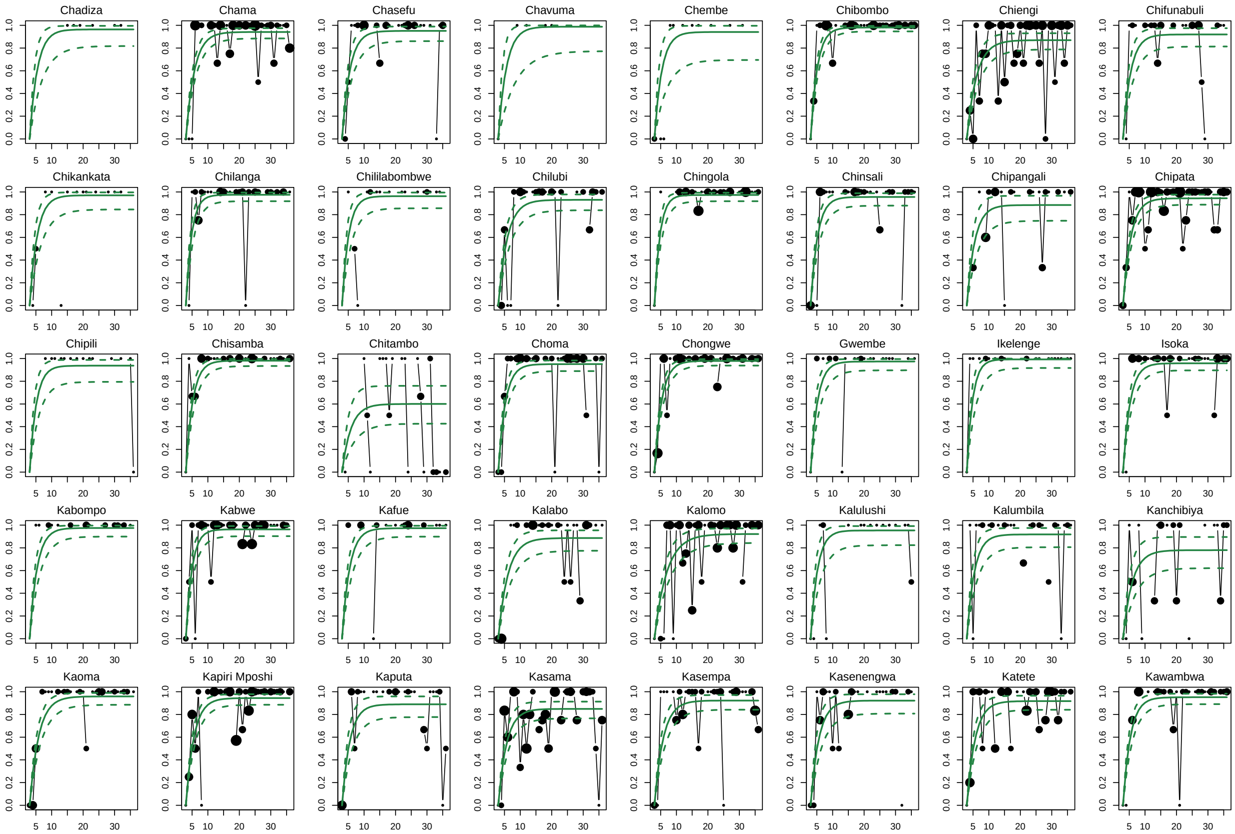
Chadiza
Chama
Chasefu
Chavuma
Chembe
Chibombo
Chiengi
Chifunabuli
Chikankata
Chilanga
Chililabombwe
Chilubi
Chingola
Chinsali
Chipangali
Chipata
Chipili
Chisamba
Chitambo
Choma
Chongwe
Gwembe
Ikelenge
Isoka
Kabompo
Kabwe
Kafue
Kalabo
Kalomo
Kalulushi
Kalumbila
Kanchibiya
Kaoma
Kapiri Mposhi
Kaputa
Kasama
Kasempa
Kasenengwa
Katete
Kawambwa

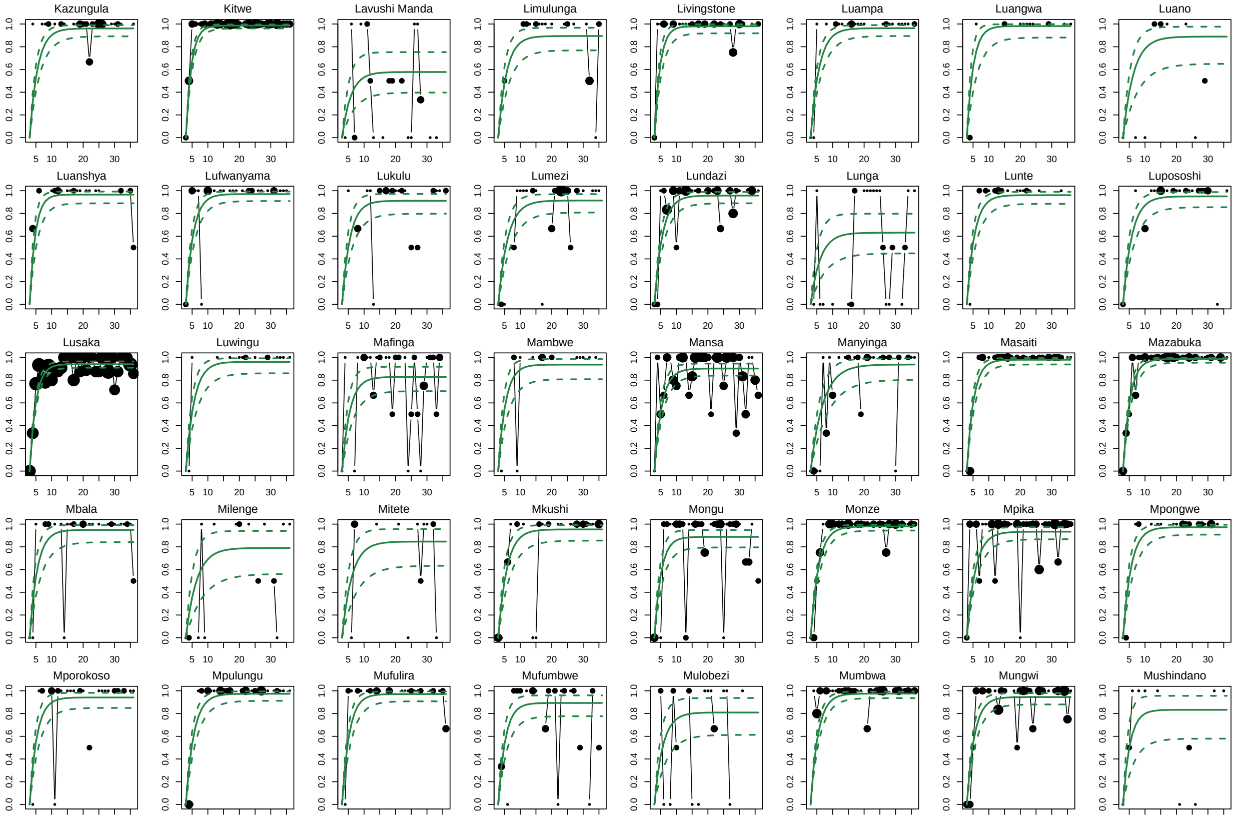
Kazungula
Kitwe
Lavushi Manda
Limulunga
Livingstone
Luampa
Luangwa
Luano
Luanshya
Lufwanyama
Lukulu
Lumezi
Lundazi
Lunga
Lunte
Lupososhi
Lusaka
Luwingu
Mafinga
Mambwe
Mansa
Manyinga
Masaiti
Mazabuka
Mbala
Milenge
Mitete
Mkushi
Mongu
Monze
Mpika
Mpongwe
Mporokoso
Mpulungu
Mufulira
Mufumbwe
Mulobezi
Mumbwa
Mungwi
Mushindano

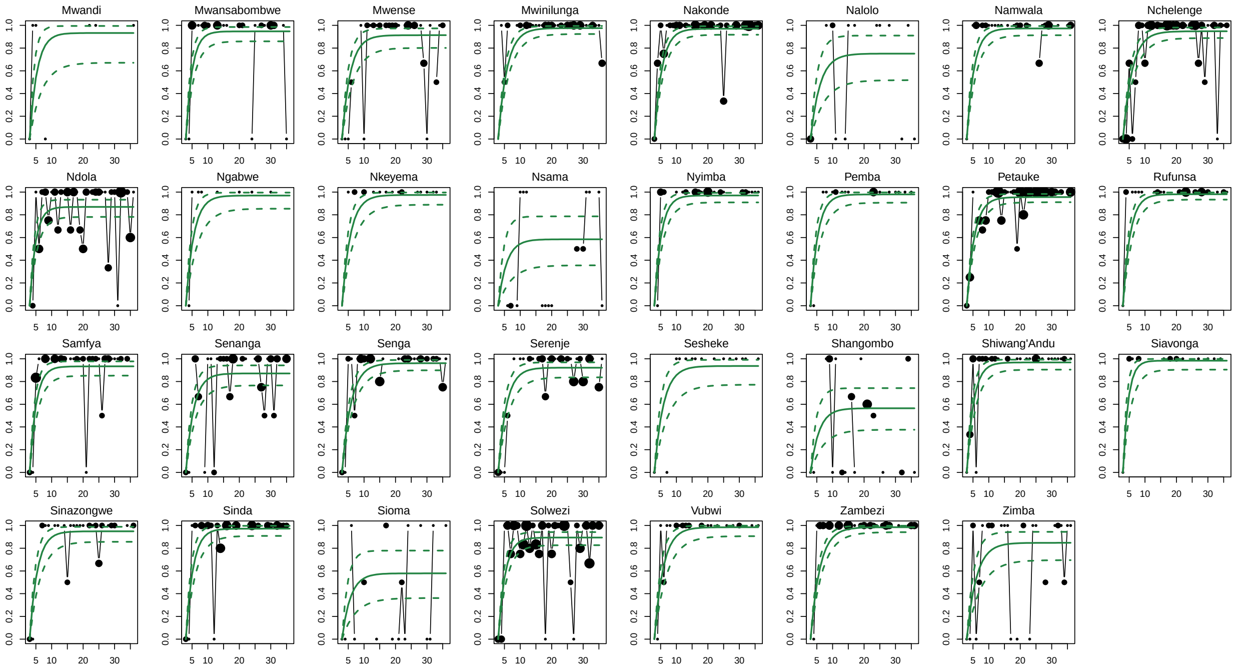


**Figure S10**: District-level Penta3 baseline routine proportion vaccinated (y-axis) over age in months (x-axis). Each plot represents one of the 111 modeled districts. The black points represent the data where the size of the point is proportional to the number of observations for each age in months. The solid and dashed lines represent the model fit median and 95% credible intervals.

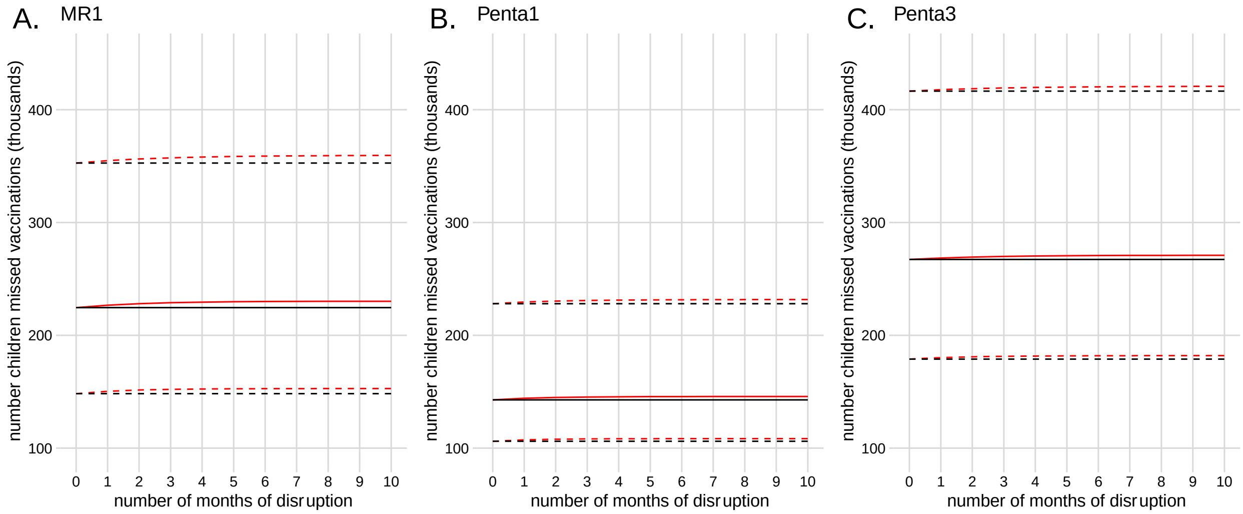


**Figure S11:** Estimated number of children missed by vaccination of MR1 (A), Penta1 (B) and Penta3 (C). The solid and dashed black lines represent the median and 95% confidence interval, respectively, of children missed by vaccination in the reference non-disruption year 2018. The solid and dashed red lines represent the median and 95% confidence interval, respectively, of children missed by vaccination in the disruption year 2020 associated with each non-disruption year percentile estimate (median and 2.5% percentile and 97.5% percentile). The estimates associated with the median number of children missed in a reference year are highlighted in Figure 3 of the main text.

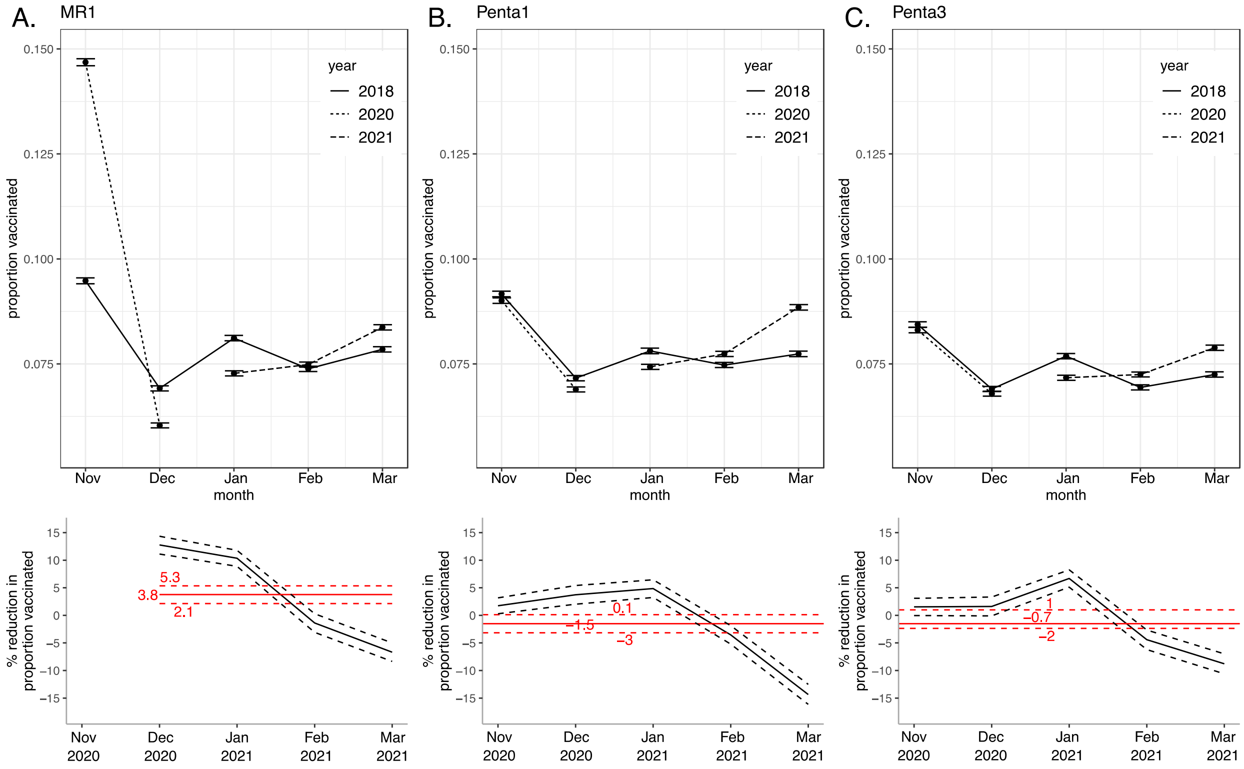


**Figure S12**: Disruption to routine vaccination for MR1 (A), Penta1 (B), Penta3 (C) based on EPI data. Top row is the proportion of the birth cohort vaccinated November and December in years 2018 and 2020 and January to March in years 2018 and 2021 (mean and 95% credible intervals represented by points and error bars). Bottom row is the percent reduction in proportion vaccinated each month (black lines) and mean across months (red lines); mean (1 - (2020/2021 mean / 2018 mean)), lower bound (1 - (2020/2021 upper bound of 95% credible interval / 2018 lower bound of 95% credible interval)), and upper bound (1 - (2020/2021 lower bound of 95% credible interval / 2018 upper bound of 95% credible interval)). The mean estimates in percent reduction of MR1 excluded November 2020 because the 2020 and 2018 are not comparable given that in 2020 an MR campaign targeting a wider age range was conducted during the child health week.
